# Disentangling phonology from phonological short-term memory in Alzheimer’s disease phenotypes

**DOI:** 10.1101/2025.01.07.25320125

**Authors:** Shalom K. Henderson, Ajay Halai, Kamen A. Tsvetanov, Thomas E. Cope, Karalyn E. Patterson, James B. Rowe, Matthew A. Lambon Ralph

## Abstract

Impaired phonological short-term memory is a core feature of the logopenic variant of primary progressive aphasia (lvPPA), but it is not clear whether a core phonological processing deficit is also present. We asked three questions: (i) beyond short-term memory impairment, do people with lvPPA have an impairment within phonology itself?; (ii) is their performance in working memory and confrontation naming reflective of this phonological impairment and/or other key contributing deficits (e.g., semantic)?; and (iii) is their repetition performance related to structural and functional differences in key language-dominant regions? We compared non- word and word repetition and short-term memory performance in patients with typical, amnestic Alzheimer’s disease (tAD, *n* = 9), lvPPA per consensus criteria (*n* = 10), and others who previously satisfied definitions of lvPPA but had progressed with multi-domain cognitive impairments (lvPPA+, *n* = 8). Bayesian analyses revealed no group differences in phonological tasks of word and non-word repetition. There was, however, a very strong group effect on multi-syllabic word/phrase repetition. We found very strong evidence for an effect of self-reported hearing loss on both word and non-word repetition, but not multi-syllabic word/phrase repetition. A comparison of phonological (as indexed by non-word repetition) *versus* working memory (as measured by multisyllabic word repetition and digit span) and confrontation naming tasks produced either no evidence or evidence for no correlation. Confrontation naming correlated positively with multisyllabic word/phrase repetition and semantic assessments across the whole group. Beyond the expected grey matter reductions in patients relative to controls in the left posterior superior temporal gyrus, anterior temporal lobe, and inferior temporal gyrus, we found no evidence for the associations between non-word repetition and functional connectivity between all regions of interest in the whole group. When excluding controls, the only anecdotal evidence that was corroborated by frequentist multiple regression was for an association between non-word repetition and functional connectivity between dorsal premotor and posterior superior temporal gyrus after controlling for the grey matter volumes in these regions. We have shown that deficits in “pure” phonological tasks, namely single word and non-word repetition, are (i) not dependent on working memory and (ii) greater in patients with a self-reported hearing loss across all groups with lvPPA or tAD. Our results suggest that instead of having a core phonological impairment, lvPPA patients have a working memory/buffering impairment that adversely affects their performance on length-dependent working memory tasks.

## 1. Introduction

The core diagnostic features of the logopenic variant of primary progressive aphasia (lvPPA) are impaired single-word retrieval and repetition of sentences and phrases, as well as secondary criteria of phonological errors in confrontation naming and speech.^1^ LvPPA is distinct from the semantic and non-fluent variants of primary progressive aphasia (underpinned by semantic and motor speech and/or grammatical impairment, respectively).^2–5^ The functional origin of deficits in lvPPA is generally attributed to impaired phonological short-term memory, sometimes used interchangeably with the term “auditory-verbal working memory”.^6–9^ The status of phonological deficits in lvPPA has been inferred previously through the use of sentence repetition, confrontation naming, and narrative samples.^10–16^

These multifaceted assessments of phonology are diagnostically informative, but there are limitations in our understanding about what they represent. First, it is unclear if lvPPA patients struggle with sentence repetition due to a core phonological processing deficit and/or an increase in working memory demands compromising their ability to repeat (i.e., hold, manipulate, and retrieve) longer word-strings and combinations.^17^ In fact, people with other PPA variants and Alzheimer’s disease (AD) fail the sentence repetition task for various reasons such as deficits in verbal working memory or motor speech.^11,18,19^ It is often assumed that deficits in short-term memory reflect the severity of phonological impairment, as it tends to be in post-stroke aphasia,^20,21^ but this has not been shown in lvPPA. Second, naming difficulties are pervasive across PPA variants and can also be found, to a lesser extent, in other subtypes of frontotemporal lobar degeneration and AD. Even though naming deficits in lvPPA are commonly thought to be due to impaired phonological processing,^14,22^ there is ongoing debate about the presence of semantic deficits in lvPPA and their potential impact on naming performance.^23–26^ Lastly, phonological paraphasias are observed in lvPPA. However, from a clinical logistics perspective, manual transcription, analysis, and quantification of phonological errors (in addition to the repairs, fillers, pauses, hesitations, and false starts that are characteristic of lvPPA speech) are unstandardised, time-consuming and impractical for clinical settings.

In the present study, we sought to address the following key questions: (i) beyond their short-term memory impairment, do individuals diagnosed with lvPPA have an impairment within phonology itself (as indexed by non-word and word repetition)?; (ii) is their performance on working memory and confrontation naming reflective of this phonological impairment and/or also other key contributing deficits (e.g., semantic)?; and (iii) is their repetition performance related to the integrity of the left hemisphere regions associated with language, in terms of either their grey matter volume or functional connectivity?

To address the first core question, one requires tasks that draw on phonological representation and processing but minimise the call upon short-term memory. In the stroke-related aphasiology literature, it is very common to use repetition tasks.^27–30^ Single word repetition reduces working memory demands relative to phrase and sentence repetition, whilst non-word repetition goes further towards a “purer” test of phonology as lexical/semantic knowledge is not needed.^21,31–35^ Only a few studies to date have assessed non-word repetition performance in individuals with lvPPA, reporting varied levels of impairment.^12,18,36^ In addition, verbal short-term memory tasks such as digit span have been found to correlate positively with sentence repetition in lvPPA,^10,19^ but how they relate to non-word repetition has not been explored. Previous studies of stroke aphasia have shown very strong parallel impairments across different types of phonological tasks including non-word repetition, reading aloud, repetition span and measures of phonological processing.^21,29,37–39^ Using phonemic discrimination and degraded speech processing tasks, prior studies of lvPPA have demonstrated the presence of a phonological input deficit that is not dependent on working memory.^40,41^ Investigating the relationship of a pure phonological test, such as single item non- word repetition, to short-term memory tests such as phrase repetition and digit span may offer valuable insights into (i) the relationship between phonological processing and working memory difficulties in lvPPA and (ii) differences in phonological profiles between post-stroke aphasia and lvPPA.

Word retrieval deficits in lvPPA have been attributed to phonological rather than other types of impairment, such as the degradation of object knowledge/semantic representation central to semantic variant PPA (svPPA)/semantic dementia (SD).^22,42–44^ Recent studies have investigated semantic cognition in lvPPA.^9,45,46^ In a previous study,^47^ we demonstrated that, although individuals with lvPPA do have semantic impairments, these are of a distinctively different type and degree to that observed in svPPA/SD. In contrast to the representational degradation in svPPA/SD and more akin to the pattern observed in semantic aphasia and Wernicke’s aphasia patients,^48–53^ lvPPA patients exhibited a deficit in semantic control (the ability to access and manipulate semantic representations flexibly for a specific context or task), showing impairments on verbal and non-verbal tests characterised by greater semantic demand.^47^ There remains a need to investigate the extent to which the word retrieval problems in lvPPA are due to their phonological impairment (to be measured via non-word repetition) and/or semantic weakness (to be measured via semantic assessments).

This experimental design also offers a valuable opportunity to investigate the neural and auditory bases of cognitive processes contributing to phonological processing and verbal short-term memory. The former issue is informed by many models of language processing and prior neuroimaging studies of healthy adults and people with post-stroke aphasia which suggest that phonological short-term memory (most frequently measured by repetition and digit span tasks) is subserved by the arcuate fasciculus connections between the left posterior superior temporal gyrus and inferior frontal gyrus.^54–58^ Previous studies have demonstrated that pseudoword repetition, in particular, engages the dorsal premotor region which is important for articulatory planning and articulation.^59–62^ This region has also been found to be specific to non-word reading in chronic stroke patients.^38^ The anterior temporal lobe is another key region that is relevant to word-, phrase- and sentence-repetition, reciprocally connected with the primary auditory cortex, and implicated in the representation and processing of semantic information.^63–67^ The growing body of evidence suggesting the link between hearing loss and dementia, particularly in individuals with AD pathology,^17,41,68^ motivated our study to assess the influence of participants’ self-reported hearing status on repetition performance.

To address our questions, we assessed performance in phonology (indexed by non-word and word repetition) *versus* short-term memory (indexed by multi-syllabic word/phrase repetition and digit span tasks) and confrontation naming, and examined imaging patterns in people with lvPPA or typical, amnestic AD as a contrast group with the same likely pathology, compared to age-matched healthy controls.

## 2. Materials and methods

### 2.1 Participants

As previously described,^47^ 27 patients diagnosed with lvPPA and AD from specialist clinics for memory disorders within the Cambridge University Hospitals and 12 age-matched healthy controls took part in this study. All participants were “White” and reported English as their first language. At the time of study participation, nine patients had a typical, amnestic presentation of AD (tAD),^69^ ten patients met strict criteria for lvPPA,^1,70^ and eight patients were classified as “lvPPA+” as they previously satisfied definitions of lvPPA but, at the time of this study, they exhibited multi-domain cognitive impairments. All participants gave written informed consent in accordance with the Declaration of Helsinki, with patients being supported by family when necessary.

Nine patients (tAD *n* = 3; lvPPA *n* = 2; lvPPA+ *n* = 4) were tested longitudinally. Cognitive assessment using the same battery (described below) was performed on two separate occasions, at the initial assessment and again at a follow-up visit. The average time between initial and follow-up was 26 months with a minimum of 18 and maximum of 36 months.

### 2.2 Assessments

Phonological tasks included word- and non-word repetition subtests from the Psycholinguistic Assessment of Language Processing in Aphasia (PALPA).^71^ Verbal working memory tasks included: (1) multisyllabic word and phrase repetition from the revised Addenbrooke’s Cognitive Examination (ACE-R),^72^ and (2) the forward and backward digit span from the Wechsler Memory Scale – Revised.^73,74^ The multisyllabic word repetition of the ACE-R includes four words (‘hippopotamus’, ‘eccentricity’, ‘unintelligible’, ‘statistician’) and we modified the standard scoring such that each word received a score of 1 or 0. The scores on these four words and two phrases of the ACE-R (i.e., ‘above, beyond, and below’ and ‘no ifs, ands or buts’) were combined to represent multisyllabic word/phrase repetition with a maximum score of six.

Confrontation naming and semantic assessments included the Boston Naming Test (BNT),^75^ including scores for spontaneously correct items and additional items correct with phonemic cues, Cambridge Semantic Battery (CSB)^2^ picture naming, the Camel and Cactus Test (CCT), and synonym judgement task,^76,77^ and the reading subcomponent of the ACE-R (i.e., reading of five words with atypical spelling-to-sound correspondences, e.g., *pint*, *sew*). Like the scoring modification previously described, each irregular word received a score of 1 or 0. Of note for later, this ACE-R reading subcomponent (i.e., exception-word reading test) is linked to semantic ability as these words are typically regularised in svPPA/SD patients due to their surface dyslexia, for example pint pronounced like “hint” and sew like “dew”.^78,79^

Picture description tasks were employed to elicit connected speech samples, involving three pictures: the Western Aphasia Battery (WAB)^80^ ‘picnic’, Boston Diagnostic Aphasia Examination (BDAE)^81^ ‘cookie theft’, and Mini Linguistic State Examination (MLSE)^82^ ‘beach’. Connected speech samples were audio recorded and transcribed by a speech-language pathologist (SKH) using the f4transcript notation software version 8.2.2, which has been previously reported to make the manual writing of speech samples from audio recordings more efficient. Speech samples were formatted for analysis with the Frequency in Language Analysis Tool.^83^ We quantified the occurrence of phonological errors during connected speech using methods reported in previous studies.^12,16,84,85^ In addition to the quantification of phonological paraphasia(s) per 100 words, we counted the number of fillers, pauses between utterances, and repaired sequences per 100 words. We defined ‘repaired sequences’ similar to Wilson *et al*.^16^ who used the following definition: “sequences of one or more complete words, which were made redundant by subsequent repetitions, amendments, elaborations or alternative expressions”. However, we included utterances in addition to complete words because repairs at the phonemic level (e.g., s- say- satin- sittin-) are representative of lvPPA speech. Given the detailed transcriptions completed, we also quantified the following measures: speech (inclusive of non-word utterances) to pause ratio, words to non-words ratio, words per minute, total utterances (including words, fillers, and non-word utterances in the repaired sequences) per minute, fillers per minute, pauses between speech segments (including non-word utterances) per minute, long pauses (lasting more than 2 seconds) per minute, short pauses (lasting 2 seconds or less) per minute, and number of repaired sequences per minute.

At the initial assessment, all participants completed the test battery except for one lvPPA+ case (Participant 1 in Supplementary Table 1) who did not complete the synonym judgement, CCT, and the ACE-R due to difficulty following task instructions. In addition, for another lvPPA+ patient (Participant 5 in Supplementary Table 1), details regarding which irregular words were read accurately for the reading subcomponent of the ACE-R were not recorded due to a technical difficulty. At the follow-up visit, testing was attempted but not completed for three patients due to severe language and cognitive impairments.

### 2.3 Neuroimaging acquisition and analysis

Twelve healthy controls participants and 27 patients completed a T1-weighted 3T structural MRI scan on a Siemens PRISMA at the University of Cambridge (GRAPPA acceleration factor = 2).^47^ Thirty-five participants were scanned at the MRC Cognition and Brain Sciences Unit with the following parameters: sagittal image acquisition, no. slices = 208, TR = 2000ms, TE = 2.85mg, flip angle = 8°, FOV = 228.8 x 281.6 x 281.6mm^3^, resolution matrix = 208 x 256 x 256, voxel size = 1.1mm^3^. Four participants were scanned at the Wolfson Brain Imaging Centre with the following parameters: sagittal image acquisition, no. slices = 208, TR = 2000ms, TE = 2.93, flip angle = 8°, FOV = 228.8 x 281.6 x 281.6mm^3^, resolution matrix = 228.8 x 256 x 256, voxel size = 1.1mm^3^.

The T1-weighted MPRAGE images were preprocessed using the processing stream of the Computational Anatomy Toolbox 12 (CAT12) (https://www.neuro.uni-jena.de/cat/) in the Statistical Parametric Mapping software (SPM12: Wellcome Trust Centre for Neuroimaging, https://www.fil.ion.ucl.ac.uk/spm/software/spm12/). Our pre-processing pipeline used: (i) denoising, resampling, bias-correction, affine registration, and brain segmentation into three tissue probability maps (grey matter, white matter, cerebrospinal fluid); (ii) normalisation and registration to the Montreal Neurological Institute (MNI) template, (iii) smoothing using 8mm full-width-half-maximum Gaussian kernel. Segmented, normalised, modulated, and smoothed grey and white matter images were used for our region of interest (ROI) analysis. Volume was quantified as the number of grey matter voxels in each ROI controlling for age and total intracranial volume as nuisance covariates.

Our *a priori* ROIs included four regions within the left language network that have been previously reported to be important for non-word reading and repetition. Ten millimetre spherical ROIs were constructed in MRIcron using the following previously published MNI coordinates: (1) posterior superior temporal gyrus (pSTG; x = -54, y = -34, z = 15)^18^; (2) inferior frontal gyrus/pars triangularis (IFG; x = -54, y = 24, z = 3)^86,87^; (3) anterior temporal lobe (ATL; x = -50, y = 11, z = -32)^87^; and (4) precentral gyrus/dorsal premotor region (x = -58, y = 4, z = 26)^38^.

Thirty-three participants also completed a resting state functional MRI scan using a T2*- weighted echoplanar sequence with the following parameters: no. slices = 42, TR = 2500ms, TE = 30ms, flip angle = 80°, FOV = 126 x 192 x 224.6mm^3^, resolution matrix = 42 x 64 x 64, voxel size = 3 x 3 x 3.5mm^3^. Participants were instructed to stay awake with eyes open during this task-free scan. Functional images were preprocessed using *fMRIPrep 23.1.4*^88,89^ which involved a standard processing pipeline including slice timing, realignment, co-registration to the T1-weighted reference, and resampling into standard space, generating a preprocessed BOLD run in MNI152NLin2009cASym space. The outputs from *fMRIPrep* were then applied to the standard functional connectivity module of the rsHRF toolbox.^90,91^ Images were smoothed at 8mm FWHM. Denoising parameters included regressing out from each time series nuisance covariates, including six motion parameters, framewise displacement outliers, first three components from the white matter and cerebrospinal fluid masks, linear detrending, and despiking. The preprocessed time series were extracted from the 10mm spherical ROIs mentioned above for our functional connectivity analysis. Time series correlations were examined between the four ROIs using Spearman’s correlations and the resulting *r* values were converted into *z*-values using the Fisher transformation.

Of the nine patients who were followed longitudinally, six completed T1-weighted 3T structural and resting state functional MRI scans using the same parameters, detailed above.

### 2.4 Statistical analysis

To test for group differences in demographics and neuropsychological performance, we conducted a Bayesian ANOVA followed by a *post hoc* test for pairwise multiple comparisons. To assess the effects of group/diagnosis, presence of hearing loss, and their interaction on repetition accuracy, we used 4 groups (i.e., controls, tAD, lvPPA, lvPPA+) by 2 hearing loss status (coded as 1 for present and 0 for absent) Bayesian ANOVA using the PALPA word, non-word, and ACE-R multisyllabic word/phrase repetition data.

Next, we used Bayesian Pearson correlations to assess the relationship between non-word and ACE-R (i.e., multisyllabic words/phrases) repetition performance and measures of (i) short-term memory: digit span forward and backward; (ii) phonology: effect of phonemic cues on the BNT (i.e., additional items correct with phonemic cues) and connected speech measures, particularly the occurrence of phonological paraphasia(s) per 100 words; and (iii) confrontation naming: BNT and CSB naming. As an exploratory analysis, we assessed the relationship between non-word and ACE-R (i.e., multisyllabic words/phrases) repetition performance and semantic tasks including the synonym judgement task, CCT, and irregular word reading from the ACE-R. Controls were excluded in these analyses as they exhibited ceiling effects.

For our imaging aims, grey matter volumes and functional connectivity *z*-values between our four ROIs were compared between controls and each patient group using a Bayesian ANOVA. Next, Bayesian linear regression analyses assessed the relationship between repetition scores (i.e., PALPA non-word, ACE-R multisyllabic word/phrase) and grey matter volumes in the left pSTG, ATL, IFG, and the dorsal premotor/precentral gyrus. To control for the potentially confounding effect of atrophy on functional connectivity, we used Bayesian multiple regression analyses to assess the relationship between repetition scores and functional connectivity *z*-values between all ROIs including the grey matter volumes in each of the relevant ROIs as covariates.

Given the relatively small number of patients who had a follow-up visit with cognitive testing and/or MRI, we employed a Bayesian approach as an exploratory analysis to test whether their cognitive performance and functional connectivity between the ROIs were significantly different from their representative diagnostic group as the normative sample across the two time-points. To be specific, the group sample data were derived from the initial assessment scores and functional connectivity measures with the sample size being 18 for lvPPA (i.e., lvPPA and lvPPA+ combined) and nine for tAD. Using the point and interval estimates of effect sizes for single case to normative sample design in neuropsychology,^92,93^ we derived a point estimate of the effect size for the difference between each patient and the group sample with an accompanying 95% credible interval, as well as a point and interval estimate of the abnormality of the patient’s score. These values indicated the percentage of the group sample that would obtain a lower score than the participant. In other words, we compared a particular participant’s cognitive scores and functional connectivity *z*-values to their diagnostic group average at initial assessment and then at follow-up visit.

All Bayesian analyses were conducted in JASP version 0.18.3.0 with secondary frequentist statistics. Evidence in favour of the alternate hypothesis was interpreted using standard thresholds with the Bayes Factor (BF): anecdotal (3 > BF > 1), moderate (10 > BF > 3), strong (30 > BF > 10), very strong (100 > BF > 30), and extreme (100 > BF). BF of less than 0.33 provided evidence for the null hypothesis. Of note, 0.33 < BF < 3 implies the lack of sufficient precision (cf. power) to draw inferences either in favour of the null or alternative hypothesis.

## 3. Results

### 3.1 Demographics and clinical details

Demographic, clinical, and neuropsychological details for each group are shown in Table 1. Bayesian ANOVA revealed evidence for no difference in all groups for age, handedness, and self-reported hearing loss status (*P* > 0.05, BF < 0.33) and there was no evidence for a difference in gender (*P* > 0.05, BF = 0.54). The results of a Bayesian ANOVA showed anecdotal evidence for a difference in self-reported symptom duration for patients (*P* = 0.05, BF = 1.66), which was driven by patients with tAD having longer symptom duration than those with lvPPA (*P* = 0.04, BF = 3.36). The anecdotal evidence for education (*P* = 0.05, BF = 1.91) was driven by controls having higher levels of education than patients, but the results of pairwise multiple comparisons did not reveal any differences between controls and each of the patient groups, and patient groups also did not differ from one another (*P* > 0.05, 0.33 < BF < 2.00).

**Table 1.**
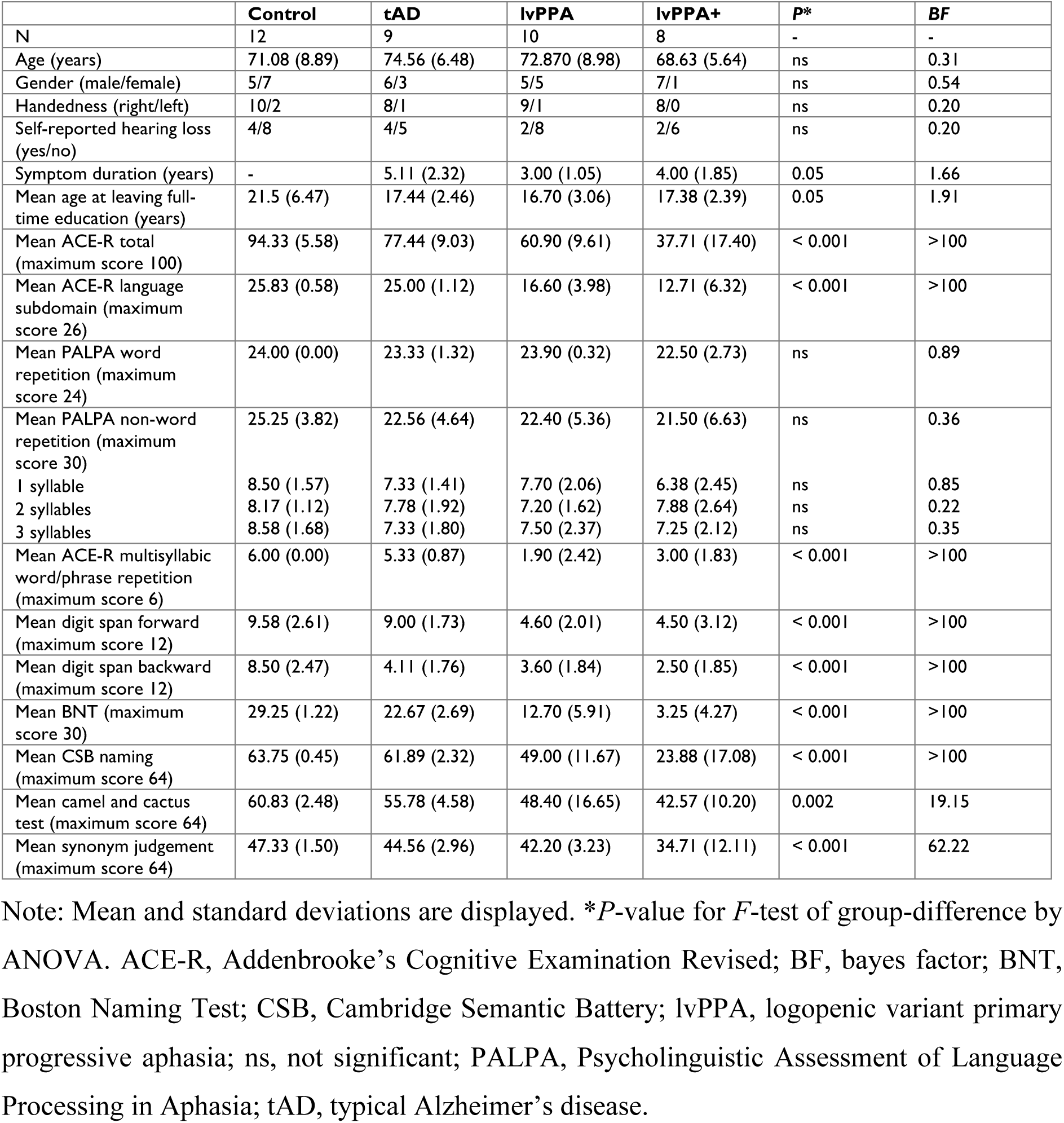
Demographics and neuropsychological data of the study cohort.

|  | Control | tAD | lvPPA | lvPPA+ | P* | BF |
| --- | --- | --- | --- | --- | --- | --- |
| N | 12 | 9 | 10 | 8 | - | - |
| Age (years) | 71.08 (8.89) | 74.56 (6.48) | 72.870 (8.98) | 68.63 (5.64) | ns | 0.31 |
| Gender (male/female) | 5/7 | 6/3 | 5/5 | 7/1 | ns | 0.54 |
| Handedness (right/left) | 10/2 | 8/1 | 9/1 | 8/0 | ns | 0.20 |
| Self-reported hearing loss (yes/no) | 4/8 | 4/5 | 2/8 | 2/6 | ns | 0.20 |
| Symptom duration (years) | - | 5.11 (2.32) | 3.00 (1.05) | 4.00 (1.85) | 0.05 | 1.66 |
| Mean age at leaving full-time education (years) | 21.5 (6.47) | 17.44 (2.46) | 16.70 (3.06) | 17.38 (2.39) | 0.05 | 1.91 |
| Mean ACE-R total (maximum score 100) | 94.33 (5.58) | 77.44 (9.03) | 60.90 (9.61) | 37.71 (17.40) | < 0.001 | >100 |
| Mean ACE-R language subdomain (maximum score 26) | 25.83 (0.58) | 25.00 (1.12) | 16.60 (3.98) | 12.71 (6.32) | < 0.001 | >100 |
| Mean PALPA word repetition (maximum score 24) | 24.00 (0.00) | 23.33 (1.32) | 23.90 (0.32) | 22.50 (2.73) | ns | 0.89 |
| Mean PALPA non-word repetition (maximum score 30) | 25.25 (3.82) | 22.56 (4.64) | 22.40 (5.36) | 21.50 (6.63) | ns | 0.36 |
| 1 syllable | 8.50 (1.57) | 7.33 (1.41) | 7.70 (2.06) | 6.38 (2.45) | ns | 0.85 |
| 2 syllables | 8.17 (1.12) | 7.78 (1.92) | 7.20 (1.62) | 7.88 (2.64) | ns | 0.22 |
| 3 syllables | 8.58 (1.68) | 7.33 (1.80) | 7.50 (2.37) | 7.25 (2.12) | ns | 0.35 |
| Mean ACE-R multisyllabic word/phrase repetition (maximum score 6) | 6.00 (0.00) | 5.33 (0.87) | 1.90 (2.42) | 3.00 (1.83) | < 0.001 | >100 |
| Mean digit span forward (maximum score 12) | 9.58 (2.61) | 9.00 (1.73) | 4.60 (2.01) | 4.50 (3.12) | < 0.001 | >100 |
| Mean digit span backward (maximum score 12) | 8.50 (2.47) | 4.11 (1.76) | 3.60 (1.84) | 2.50 (1.85) | < 0.001 | >100 |
| Mean BNT (maximum score 30) | 29.25 (1.22) | 22.67 (2.69) | 12.70 (5.91) | 3.25 (4.27) | < 0.001 | >100 |
| Mean CSB naming (maximum score 64) | 63.75 (0.45) | 61.89 (2.32) | 49.00 (11.67) | 23.88 (17.08) | < 0.001 | >100 |
| Mean camel and cactus test (maximum score 64) | 60.83 (2.48) | 55.78 (4.58) | 48.40 (16.65) | 42.57 (10.20) | 0.002 | 19.15 |
| Mean synonym judgement (maximum score 64) | 47.33 (1.50) | 44.56 (2.96) | 42.20 (3.23) | 34.71 (12.11) | < 0.001 | 62.22 |

Note: Mean and standard deviations are displayed. \* $P$ -value for $F$ -test of group-difference by ANOVA. ACE-R, Addenbrooke's Cognitive Examination Revised; BF, bayes factor; BNT, Boston Naming Test; CSB, Cambridge Semantic Battery; lvPPA, logopenic variant primary progressive aphasia; ns, not significant; PALPA, Psycholinguistic Assessment of Language Processing in Aphasia; tAD, typical Alzheimer's disease.

As expected, evidence for a difference in ACE-R total scores between controls and all three patient groups was extreme (*P* ≤ 0.005, BF > 100). Across the patient groups, evidence for a difference in ACE-R total scores was extreme between tAD and lvPPA+ (*P* < 0.001, BF > 100), and strong between tAD and lvPPA (*P* = 0.007, BF = 24.72) and lvPPA and lvPPA+ (*P* < 0.001, BF = 12.69). For ACE-R language sub-scores, there was extreme evidence for a difference between controls and tAD versus lvPPA and lvPPA+ (*P* < 0.001, BF > 100).

There was no evidence for a difference in word and non-word repetition scores across all groups, as well as between 1 and 3 syllable length non-words (*P* > 0.05, 0.33 < BF < 1.00). We found evidence for no difference across groups for 2 syllable length non-words (*P* > 0.05, BF = 0.22). For ACE-R multisyllabic word/phrase repetition, evidence for a difference was extreme for controls versus lvPPA and lvPPA+ (*P* ≤ 0.001, BF > 100), as well as strong between tAD and lvPPA (*P* < 0.001, BF = 32.48) and moderate between tAD and lvPPA+ (*P* = 0.02, BF = 9.54). For digit span forward, evidence for a difference was extreme between controls and lvPPA (*P* < 0.001, BF > 100), as well as tAD and lvPPA (*P* = 0.002, BF > 100), very strong between controls and lvPPA+ (*P* < 0.001, BF = 31.35), and strong between tAD and lvPPA+ (*P* = 0.003, BF = 17.38). There was no evidence for a difference between controls and tAD (*P* = 0.95, BF = 0.44), as well as between lvPPA and lvPPA+ (*P* = 1.00, BF = 0.41). For digit span backwards, evidence for a difference was extreme for controls versus lvPPA+ and lvPPA (*P* < 0.001, BF > 100), and very strong versus tAD (*P* < 0.001, BF = 98.30). All other group comparisons showed anecdotal or no evidence (*P* > 0.05, 0.33 < BF < 2.00). For a deep dive into the phonological errors made by patients and controls during the non-word repetition task, see Supplementary Tables 1 and 2, respectively.

As expected, evidence for a difference in BNT scores between controls and all three patient groups was extreme (*P* < 0.01, BF > 100). Across the patient groups, evidence for a difference in BNT scores was extreme between tAD and lvPPA+ (*P* < 0.001, BF > 100), very strong between tAD and lvPPA (*P* < 0.001, BF = 95.77), and strong between lvPPA and lvPPA+ (*P* < 0.001, BF = 20.50). Similarly, evidence for a difference in CSB naming scores was extreme for controls and tAD versus lvPPA+ (*P* < 0.001, BF > 100), very strong between controls versus lvPPA (*P* = 0.006, BF = 85.09), strong between lvPPA and lvPPA+ (*P* < 0.001, BF = 17.80), and moderate between tAD and lvPPA (*P* = 0.03, BF = 8.99). For CCT, evidence for a difference was extreme between controls and lvPPA+ (*P* = 0.003, BF > 100), strong between tAD and lvPPA+ (*P* = 0.06, BF = 10.86), and moderate between controls and lvPPA (*P* = 0.03, BF = 3.33). For synonym judgement task, there was strong evidence that scores differed between controls and lvPPA+ (*P* < 0.001, BF = 16.86). Evidence was anecdotal only for lvPPA+ versus tAD (*P* = 0.007, BF = 2.31) and lvPPA (*P* = 0.05, BF = 1.31).

### 3.2 Effect of hearing loss on repetition

Results of a Bayesian group x hearing loss ANOVA revealed moderate evidence for an effect of self-reported hearing status (F(1,31) = 14.42, *P* < 0.001, BF = 9.46) and group (F(3,31) = 10.10, *P* < 0.001, BF = 4.90), as well as strong evidence for a hearing-by-group interaction (F(3,31) = 8.04, *P* < 0.001, BF = 11.84) on word repetition. Given the strong evidence for the hearing-by-group interaction, *post-hoc* Bayesian independent samples *t-*tests were conducted and revealed moderate evidence for a difference between those with and without hearing loss in the lvPPA+ group only (*t* = 3.18, *P* = 0.02, BF = 3.25); there was no evidence for a difference in the tAD group (*t* = 1.22, *P* = 0.26, BF = 0.77) and controls and lvPPA patients were mostly at ceiling resulting in zero variance for the *t*-tests.

For non-word repetition, Bayesian group x hearing loss ANOVA revealed very strong evidence for an effect of self-reported hearing status only (F(1,31) = 18.25, *P* < 0.001, BF = 49.30); there was no evidence for a hearing-by-group interaction (*P* = 0.21, BF = 0.83). There was no evidence for an effect of hearing status or hearing-by-group interaction (0.33 < BF < 1) on ACE-R multisyllabic word/phrase repetition.

### 3.3 Correlations between repetition and other tasks

As shown in Figure 2A, there was no evidence for a correlation between non-word repetition and forward digit span in the whole group (*r* = 0.29, *P* = 0.14, BF = 0.68), as well as in the individual groups: tAD (*r* = 0.42, *P* = 0.26, BF = 0.72), lvPPA (*r* = 0.12, *P* = 0.74, BF = 0.41), lvPPA+ (*r* = 0.50, *P* = 0.21, BF = 0.86), and lvPPA/lvPPA+ combined (*r* = 0.34, *P* = 0.17, BF = 0.70). There was evidence for no correlation between non-word repetition and backward digit span in the whole group (*r* = 0.09, *P* = 0.66, BF = 0.26) and in the lvPPA/lvPPA+ combined group (*r* = 0.09, *P* = 0.72, BF = 0.31). There was no evidence for a correlation between non- word repetition and backward digit span in the individual patient groups (0.33 < BF < 1). Figure 2B shows the moderate evidence found between non-word repetition and the effect of phonemic cueing on BNT (i.e., raw scores of additional items correct with phonemic cues) in the lvPPA/lvPPA+ combined group (*r* = 0.57, *P* = 0.01, BF = 5.11). This correlation was not observed in the other groups (0.33 < BF < 2). Using connected speech measures, we found moderate evidence for a correlation between non-word repetition and the number of phonological paraphasias per 100 words in the lvPPA+ group (*r* = -0.80, *P* = 0.02, BF = 4.64). There was either anecdotal or no evidence in the other groups (0.33 < BF < 3) as shown in Figure 2C.

**Figure 1.**
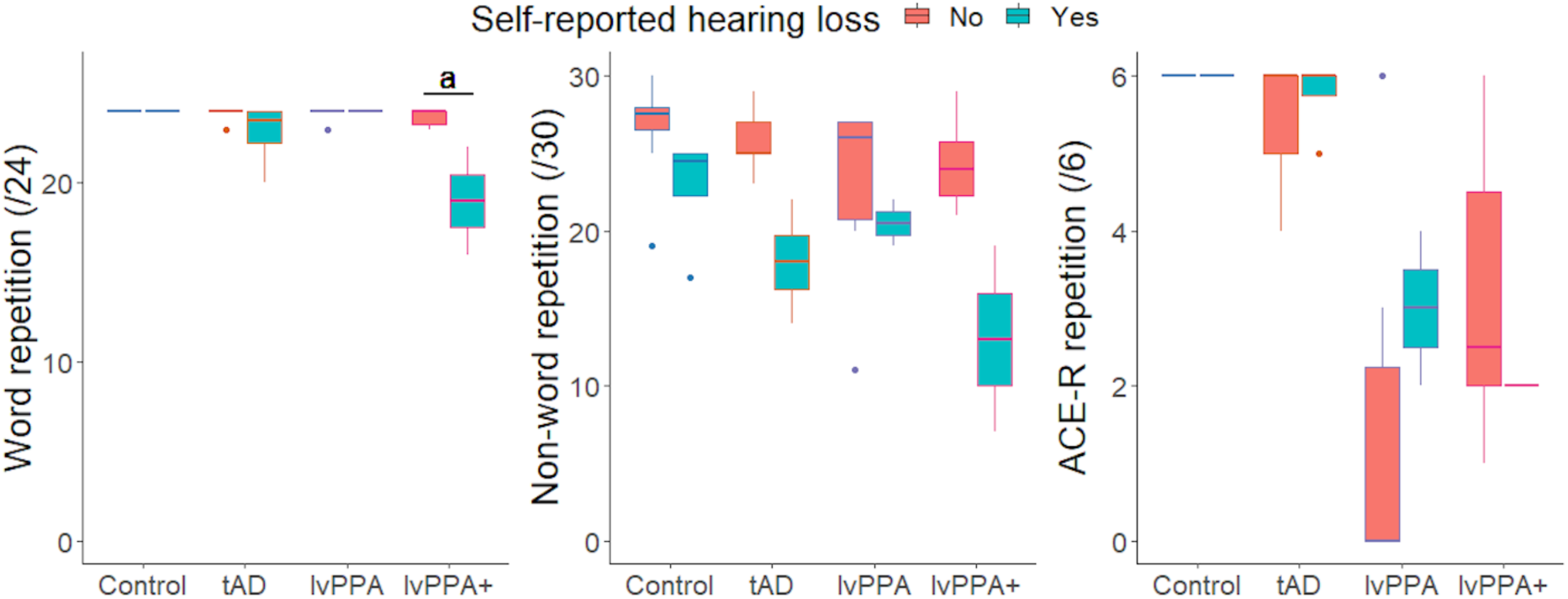
E**f**fect **of self-reported hearing loss on word (left), non-word (middle) and multisyllabic word/phrase (right) repetition performance.** Bayesian group x hearing loss ANOVA revealed moderate evidence for an effect of self-reported hearing status (F(1,31) = 14.42, *P* < 0.001, BF = 9.46) and group (F(3,31) = 10.10, *P* < 0.001, BF = 4.90), and strong evidence for a hearing-by-group interaction (F(3,31) = 8.04, *P* < 0.001, BF = 11.84) on word repetition. *Post hoc* Bayesian *t*-test revealed moderate evidence for a difference in those with and others without hearing loss in the lvPPA+ group only (*t* = 3.18, *P* = 0.02, BF = 3.25). The letter ‘a’ indicates the level of Bayesian evidence: a = moderate (3 < BF < 10). For non-word repetition, Bayesian group x hearing loss ANOVA revealed very strong evidence for an effect of self-reported hearing status only (F(1,31) = 18.25, *P* < 0.001, BF = 49.30). For ACE-R multisyllabic word/phrase repetition, there was no evidence for an effect of hearing status or hearing-by-group interaction (0.33 < BF < 1). BF, Bayes factor; lvPPA, logopenic variant primary progressive aphasia; tAD, typical Alzheimer’s disease.

**Figure 2.**
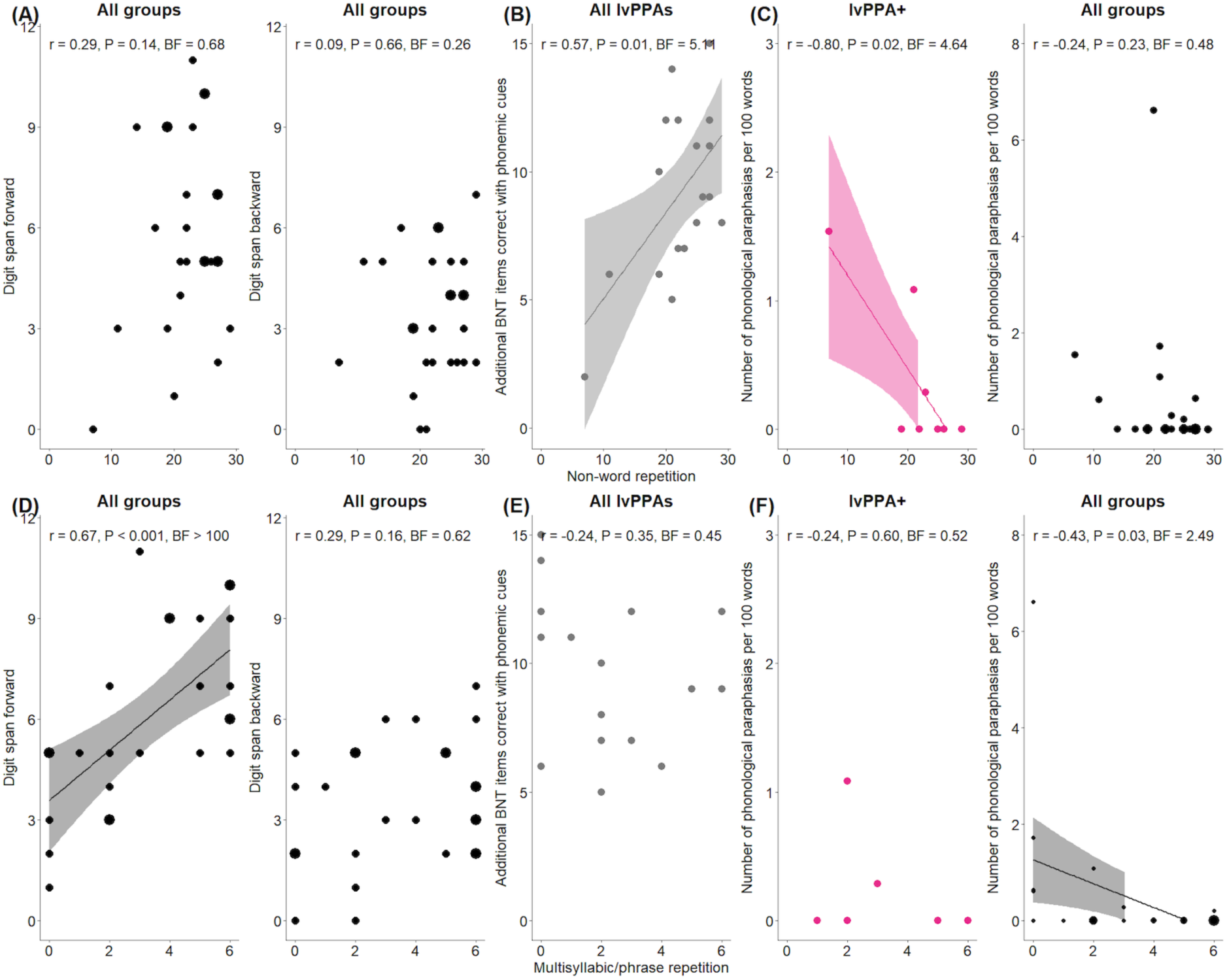
A**s**sociations **between PALPA non-word and ACE-R multisyllabic word/phrase repetition and measures of phonology and verbal working memory.** Panels indicate correlations between (**A**) non-word repetition and digit span forward (*left*) and backward (*right*) in the whole group, (**B**) non-word repetition and the additional items correct with phonemic cues on the Boston Naming Test (BNT) in the lvPPA/lvPPA+ combined group, (**C**) non-word repetition and the number of phonological paraphasias per 100 words during connected speech in the lvPPA+ group (*left*) and all groups (*right*), (**D**) ACE-R multisyllabic word/phrase word repetition and digit span forward (*left*) and backward (*right*) in the whole group, (**E**) ACE-R multisyllabic word/phrase word repetition and the additional items correct with phonemic cues on the BNT in the lvPPA/lvPPA+ combined group, and (**F**) ACE-R multisyllabic word/phrase word repetition and the number of phonological paraphasias per 100 words during connected speech in the lvPPA+ group (*left*) and all groups (*right*). In the scatterplots for the whole group, the dots indicate the number of participants (e.g., larger dots portray more participants who have the same scores). In each scatterplot, the Bayesian Pearson’s correlation values, their associated Bayes factors (BFs), and Frequentist *P* values are shown. ACE-R, Addenbrooke’s Cognitive Examination – Revised; lvPPA, logopenic variant primary progressive aphasia.

Figure 2D shows the correlations between the ACE-R multisyllabic word/phrase repetition and the same phonology and working memory tests. There was very strong evidence for a correlation between ACE-R multisyllabic word/phrase repetition and forward digit span in the whole group (*r* = 0.67, *P* < 0.001, BF > 100), as well as moderate evidence in the lvPPA group (*r* = 0.73, *P* = 0.02, BF = 4.89) and anecdotal evidence in the lvPPA/lvPPA+ combined group (*r* = 0.49, *P* = 0.04, BF = 1.95). This correlation was not observed in the tAD and lvPPA+ groups (0.33 < BF < 1). There was no evidence for correlation between ACE-R multisyllabic word/phrase repetition and backward digit span in the whole group (*r* = 0.29, *P* = 0.16, BF = 0.62), as well as in the individual patient groups (0.33 < BF < 1). There was moderate evidence for a correlation between ACE-R multisyllabic word/phrase repetition and additional items correctly named with phonemic cues on the BNT in the whole group only (*r* = -0.53, *P* = 0.006, BF = 9.22), and there was no evidence in the other groups (0.33 < BF < 2). Lastly, evidence for a correlation between ACE-R multisyllabic word/phrase repetition and the number of phonological paraphasias per 100 words during connected speech was anecdotal in the whole group (*r* = -0.43, *P* = 0.3, BF = 2.49) only. There was either anecdotal or no evidence in the other groups (0.33 < BF < 2). Supplementary Table 3 shows the correlations between all connected speech measures and repetition (i.e., PALPA non-word, ACE-R multisyllabic word/phrase) performance across groups.

In the whole group, there was evidence for no correlation between non-word repetition and CSB naming and BNT (BF < 0.33). Evidence for a correlation between ACE-R multisyllabic word/phrase repetition and BNT was strong (*r* = 0.54, *P* = 0.004, BF = 11.15) and moderate for CSB naming (*r* = 0.47, *P* = 0.02, BF = 3.79). This correlation remained significant with moderate evidence in the lvPPA+ group for CSB naming only (*r* = 0.85, *P* = 0.02, BF = 4.89). Bayesian Pearson’s correlations between PALPA non-word and ACE-R multisyllabic word/phrase repetition and semantic assessments are visually summarised in Supplementary Figure 1, with associated Bayes factors.

### 3.4 Neuroimaging

Group differences in grey matter volumes and functional connectivity between each of our ROIs are shown in Table 2. Evidence for grey matter intensity in the left pSTG being greater in controls was extreme relative to tAD and lvPPA (BF >100), and very strong relative to lvPPA+ (BF 36.84), but there was no evidence for a difference between the patient groups (0.33 < BF < 1). For the left IFG, the difference was very strong for controls versus tAD and lvPPA (BF > 30). There was either anecdotal or no evidence in the other group comparisons (0.33 < BF < 3). Evidence for grey matter intensity in the left ATL being greater in controls was extreme relative to lvPPA and tAD (BF >100), and very strong relative to lvPPA+ (BF = 48.89), but there was either anecdotal or no evidence for a difference between the patient groups (0.33 < BF < 3). For the left precentral gyrus/dorsal premotor (dPM) region, there was either anecdotal or no evidence for all group comparisons (0.33 < BF < 3). Group comparisons with resting state functional connectivity in our four voxel-based spherical ROIs revealed the following: (1) there was no evidence for difference between controls and patient groups in pSTG-dPM, and IFG-dPM functional connectivity (0.33 < BF < 1); and (2) there was evidence for no difference in functional connectivity between pSTG-IFG, pSTG-ATL, IFG-ATL, and ATL-dPM regions across all groups (BF < 0.33).

**Table 2.** Grey matter volumes and Fisher’s z-transformed ROI-to-ROI resting state functional connectivity values.

|  | Control | tAD | lvPPA | lvPPA+ | P* | BF |
| --- | --- | --- | --- | --- | --- | --- |
| **Grey matter volume in the four ROIs** |  |  |  |  |  |  |
| N | 12 | 9 | 10 | 8 | - | - |
| Left pSTG | 262.90 (31.35) | 204.13 (15.41) | 196.68 (36.92) | 207.18 (28.28) | < 0.001 | >100 |
| Left IFG | 264.47 (30.28) | 207.80 (30.77) | 209.45 (34.81) | 246.31 (45.30) | < 0.001 | 36.74 |
| Left ATL | 303.53 (36.25) | 235.52 (29.90) | 206.49 (22.76) | 224.42 (48.05) | < 0.001 | >100 |
| Left precentral/dorsal premotor (dPM) | 91.50 (10.44) | 83.14 (8.93) | 79.11 (15.82) | 87.05 (15.08) | ns | 0.72 |
| **Pairwise resting state functional connectivity z(r) values** |  |  |  |  |  |  |
| N | 12 | 6 | 9 | 6 | - | - |
| pSTG-IFG | 0.27 (0.16) | 0.27 (0.27) | 0.31 (0.21) | 0.43 (0.32) | ns | 0.27 |
| pSTG-ATL | 0.15 (0.22) | 0.12 (0.33) | 0.23 (0.12) | 0.17 (0.14) | ns | 0.21 |
| pSTG-dPM | 0.55 (0.19) | 0.36 (0.19) | 0.44 (0.20) | 0.43 (0.24) | ns | 0.48 |
| IFG-ATL | 0.33 (0.21) | 0.36 (0.10) | 0.38 (0.23) | 0.34 (0.18) | ns | 0.17 |
| IFG-dPM | 0.34 (0.17) | 0.22 (0.22) | 0.35 (0.24) | 0.47 (0.17) | ns | 0.49 |
| ATL-dPM | 0.17 (0.20) | 0.06 (0.28) | 0.07 (0.18) | 0.12 (0.11) | ns | 0.29 |

Note: Mean and standard deviations are displayed. \*P-value for *F*-test of group-difference by ANOVA. Grey matter volumes were extracted for all participants and thirty-three participants (85% of total sample) completed the resting state fMRI scan. The column entitled 'N' reflects these numbers. ATL, anterior temporal lobe; BF, Bayes factor; dPM, dorsal premotor region; IFG, inferior temporal gyrus; lvPPA, logopenic variant primary progressive aphasia; ns, not significant; pSTG, posterior superior temporal gyrus; tAD, typical Alzheimer's disease.

In the whole group, we found evidence for no relationship between non-word repetition and IFG grey matter volumes (BF = 0.31). In addition, we found no evidence for a relationship between non-word repetition scores and grey matter volumes in the other ROIs, as well as connectivity between all ROIs. As shown in Figure 3A, the results of Bayesian linear regression analysis revealed evidence for the associations between ACE-R multisyllabic word/phrase repetition and grey matter volumes in the pSTG (*β* = 0.58, *t* = 4.22, *P* < 0.001, BF > 100), ATL (*β* = 0.58, *t* = 4.26, *P* < 0.001, BF > 100), and IFG (*β* = 0.39, *t* = 2.52, *P* = 0.02, BF = 3.44).

**Figure 3.**
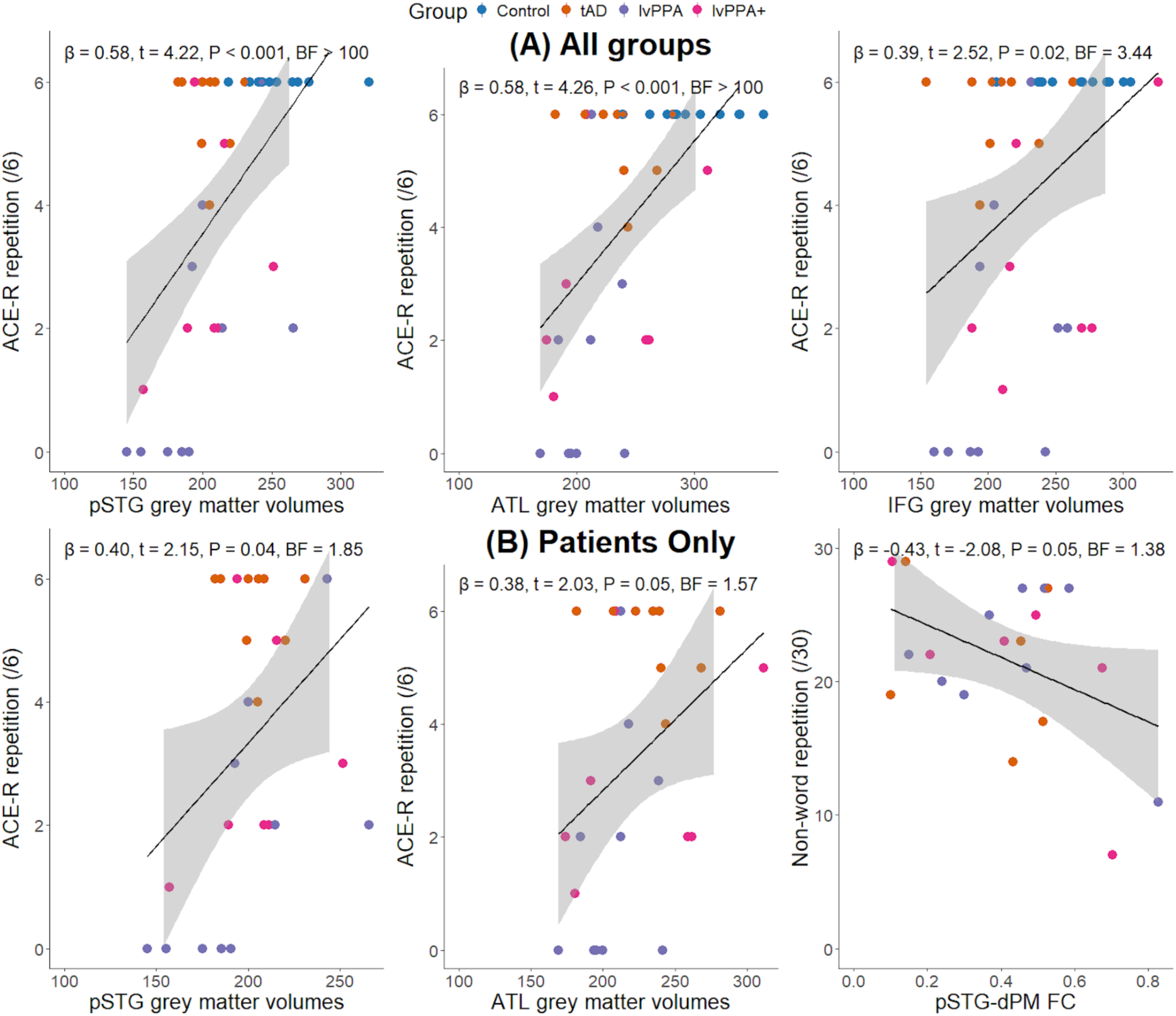
A**s**sociations **between non-word and multisyllabic word/phrase repetition scores and region of interest (ROI) grey matter volumes and functional connectivity**. Panel (**A**) shows the association between ACE-R multisyllabic word/phrase repetition and grey matter volumes in the left posterior superior temporal gyrus (pSTG; *left*), anterior temporal lobe (ATL; *middle*), and IFG (*right*) in the whole group. Panel (**B**) shows the association between ACE-R multisyllabic word/phrase repetition and grey matter volumes in pSTG (*left*), ATL (*middle*), and the association between non-word repetition and pSTG-dorsal premotor (dPM) resting state functional connectivity after controlling for grey matter volumes in the pSTG and dPM (*right*) in patients only. Each scatterplot shows the results of a Bayesian multiple regression, including the beta coefficients, associated Bayes factors (BFs), and Frequentist *P* values. ACE- R, Addenbrooke’s Cognitive Examination – Revised; FC, functional connectivity.

Evidence for the association between ACE-R multisyllabic word/phrase repetition and grey matter volumes in the dPM region was anecdotal (*β* = 0.29, *t* = 1.79, *P* = 0.08, BF = 1.08). Bayesian multiple regression analysis revealed anecdotal or no evidence for the associations between ACE-R multisyllabic word/phrase repetition and functional connectivity in all ROIs after controlling for the grey matter volumes in the relevant ROIs (0.33 < BF < 3).

When excluding controls, we found no evidence for a relationship between non-word repetition scores and grey matter volumes in all ROIs and functional connectivity (0.33 < BF < 1). The only anecdotal evidence that was corroborated by frequentist multiple regression was for an association between non-word repetition and pSTG-dPM functional connectivity (*β* = -0.43, *t* = -2.08, *P* = 0.05, BF = 1.38) after controlling for the grey matter volumes in the pSTG and dPM. As shown in Figure 3B, we found anecdotal evidence for the associations between ACE- R multisyllabic word/phrase repetition and grey matter volumes in the pSTG (*β* = 0.40, *t* = 2.15, *P* = 0.04, BF = 1.85) and ATL (*β* = 0.38, *t* = 2.03, *P* = 0.05, BF = 1.57). Bayesian multiple regression analysis revealed anecdotal (0.33 < BF ≤ 1.4) or no (0.33 < BF <1) associations between ACE-R multisyllabic word/phrase repetition and functional connectivity in all ROIs.

### 3.5 Longitudinal change

Bayesian comparisons between each longitudinal case and their representative diagnostic group sample are shown in Supplementary Table 4. Figure 4 shows the performance on repetition, digit span and BNT over time in six patients who completed the neuropsychological battery at two time-points. Participant 5, who was classified as lvPPA+ and most impaired on repetition at initial assessment, showed the greatest decline in both word and non-word repetition over time. The two-tailed probability of < 0.001 and 0.008 for word and non-word repetition, respectively, indicates that the patient’s score was significantly (*P* < 0.05) below the lvPPA group mean and that an estimated 0% – 0.40% of the entire lvPPA sample would obtain a score lower than the patient’s. Participant 5 also exhibited a significant decline in semantics over time as shown in the two-tailed probability of 0.03 and a Bayesian estimated percentage of 1.49% for CCT at the second time-point. As shown in Figure 4, the two lvPPA patients (Participants 18 and 22) showed declines in all tests except for word repetition. Of the three tAD patients, only Participant 24 showed decline in all three repetition tasks over time, including a significant decline in word repetition with a two-tailed probability of 0.04 and a Bayesian estimated percentage of 1.78%. Another tAD patient (Participant 19) exhibited significant declines in naming and semantics, as shown in the two-tailed probabilities and Bayesian point estimates for the BNT, CSB naming, and CCT. As shown in Supplementary Figure 2, there was considerable variability between functional connectivity *z* -values between patients, as well as in the pattern of change over time. Except for Participant 22 having significantly higher IFG-ATL FC *z*-values at the first time-point relative to the tAD group sample, none of the FC values showed two-tailed significant between first and second time- points in the six patients as shown in Supplementary Table 4.

**Figure 4.**
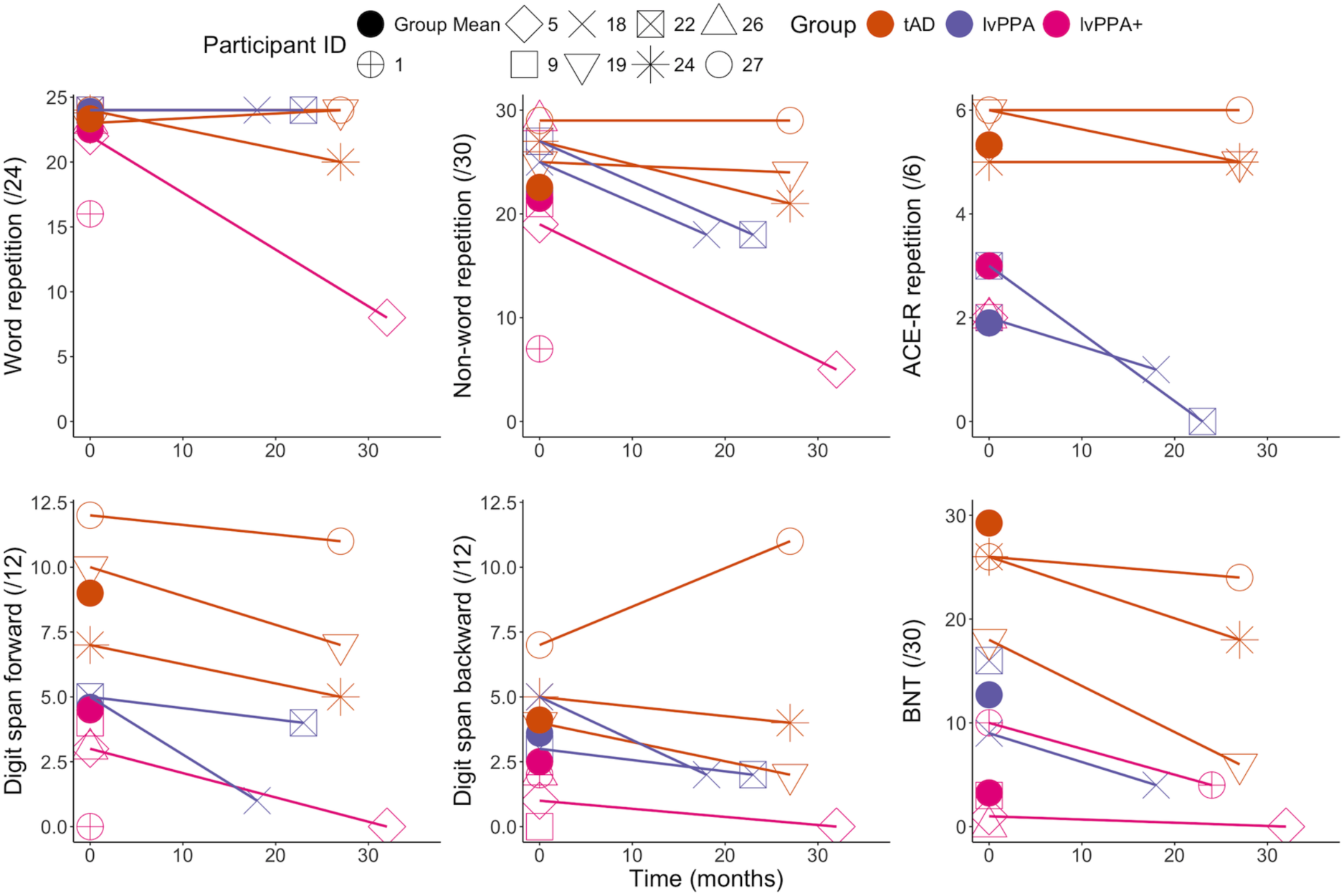
C**o**gnitive **scores over time in six patient participants**. The x-axis represents the time from to initial to follow-up assessment in months. The filled colour dots represent the group average scores on each test. The connected lines indicate the scores of each participant who completed the same test at two time-points. Note: The lines merely connect the same participants across two time-points and do not suggest a linear decline over time.

## Discussion

Impaired phonological short-term memory is the primary diagnostic feature of lvPPA^6–9^ and verbal span tasks like sentence repetition are widely used as the “gold standard” assessment to detect this core impairment.^7,10,13^ However, it was unclear if lvPPA patients struggle with sentence repetition due to a “pure” phonological processing deficit (i.e., to the phonological representations themselves) and/or short-term memory impairment. Secondly, even though word retrieval deficits – one of the two core features of lvPPA – are generally assumed to be due to impaired phonological rather than other deficits (e.g., the object knowledge/semantic representation degradation central to svPPA/SD), the influence of semantic deficits on naming performance needs further investigation given the growing body of evidence showing impaired semantic performance in lvPPA.^9,45,46^ Lastly, while there are phonological paraphasias or paraphasic errors in lvPPA speech, formal quantification of such errors is rare; and how phonological paraphasias relate to phonological tasks, including “purer” tests such as non-word repetition, remains largely unexplored. In the present study, we sought to address the following key questions: (i) beyond their short-term memory impairment, do individuals diagnosed with lvPPA have an impairment within phonology itself (as indexed by single non-word and word repetition)?; (ii) is their performance on working memory and naming reflective of this phonological impairment and/or also other key contributing deficits (e.g., semantic)?; and (iii) is their repetition performance related to (a) the grey matter volumes in the key language-dominant left hemisphere regions that are associated with phonological processing, and (b) to the functional connectivity between these regions? In the following sections, we revisit these questions, interpret our findings, and note directions for future research.

### Do lvPPA patients have a core phonological impairment?

Only a few studies to date have reported non- or pseudo-word repetition performance in individuals with lvPPA.^7,18,36^ Similar to their findings, performance across patients in our study was variable, with some lvPPA (Participants 1 and 2) and tAD (Participants 3 and 4) patients making numerous errors whereas other lvPPA (Participants 25 and 26) and tAD (Participants 24 and 27) cases exhibited ceiling effects (see Supplementary Table 1).

There are three main findings that highlight striking differences between this study of lvPPA compared with cases in prior reports. Here, we focus on the post-stroke aphasia literature given the paucity of lvPPA studies that have assessed non-word repetition. Firstly, there was no evidence for an effect of group on word and non-word repetition; in other words, all patient groups performed similarly on these phonological tests. Significant breakdowns, however, occurred in the lvPPA and lvPPA+ groups when word length increased (greater than 3 syllables, e.g., ‘hippopotamus’) and when phrase length increased (e.g., ‘no ifs, ands, or buts’). This uniquely differentiates patients with lvPPA from previously reported post-stroke aphasics for whom non-word repetition was found to be highly correlated with their performance across all types of repetition stimuli.^21,29^

Second, performance on non-word repetition in the whole and individual patient groups did not correlate with ACE-R multisyllabic word/phrase repetition or digit span tasks (both forward and backward). In contrast, phonological problems from prior studies of stroke aphasia include strong positive correlations across phonological tasks, between phonological and working memory tasks, as well as between phonological abilities and the frequency of phonemic errors.^20,29,37,38^ Lexicalisation errors have also been found to be frequent in post-stroke aphasics and provide another contrast between lvPPA and post-stroke cases.^29,35,94^ These findings suggest that, by contrasting single item repetition to longer, multi-item span, we can pull apart differences between phonology itself and phonological short-term memory. Unlike the post-stroke aphasic cases, lvPPA patient exhibited relatively preserved phonology but impaired short-term memory. Recent studies utilising degraded speech processing and phonemic discrimination tasks have shown that individuals with lvPPA demonstrate a phonological input deficit that is not dependent on working memory,^40,41^ suggesting that the status of phonological impairment in lvPPA may be modulated by task and context (much like semantic control reported in our previous study^47^). While the hallmark of lvPPA is breakdown in phonological short-term memory and the integrity of phonological representations, the effects of degraded phonological representations on the control system (e.g., struggling to discriminate, assemble, or monitor speech sounds accurately) need further investigation.

Lastly and importantly, we found evidence for an effect of self-reported hearing status on word and non-word repetition, but not on multisyllabic word/phase repetition. In other words, participants (including controls) who reported to have a hearing loss performed significantly worse on word and non-word but not on longer repetition tasks with greater working memory demands. Our findings raise a further question: does the presence of self-reported hearing loss account for non-word repetition deficits or exacerbate the underlying phonological impairment? For word repetition, we found evidence for an effect of hearing loss and hearing- by-group interaction. *Post hoc* analysis revealed that only lvPPA+ patients with a hearing loss were more impaired when compared to patients without a hearing loss. For non-word repetition, we did not find this interaction; in other words, regardless of group, all controls and patients performed more poorly on repeating non-words if they had a hearing loss. One possible interpretation is that controls and patients with tAD and lvPPA can overcome auditory input problems by utilising top-down information to repair phonological output with the use of lexical-semantics when the words are real, but this process of repair becomes difficult when the words are not real. This type of suppressed phonemic restoration has been found in AD patients by Jiang *et al*.^95^ who proposed that the largely preserved top-down semantic mechanisms (cf. impaired auditory parsing and phonemic representations) override the repair of real words. It is also possible that phonological deficits may lead to a symptom that is described as a hearing loss. Previous studies have suggested that early brain changes associated with neurodegenerative diseases such as AD might affect the auditory brain network which manifest as symptoms of hearing loss that are not detectable with standard hearing assessments,^17,96–98^ and thus the link between central hearing and phonological impairment in lvPPA requires further investigation. When excluding those with a self-reported hearing loss, most lvPPA and lvPPA+ patients showed intact performance on word and non-word repetition. This finding suggests that, instead of having a core phonological impairment, lvPPA patients may have a buffering information-type impairment.^99–102^

In summary, the patterns of word and non-word performance and the lack of a correlation between non-word repetition and other canonical phonological short-term memory tasks suggests that lvPPA patients (1) deviate from the stroke aphasia model (as explained above), and (2) have relatively intact phonological representations but impaired short-term memory/information buffering.

### Is naming influenced by phonology and/or semantics in lvPPA?

Anomia is a common feature shared across PPA variants, as well as among typical and atypical subtypes of AD. While confrontation naming tests are excellent at detecting the presence and degree of word retrieval deficits, there is no agreement on *why* individuals with lvPPA are impaired. The commonalities among the many models of word retrieval and speech production include four broad stages in naming: visual recognition, semantic activation, phonological activation, and speech production. After an image (e.g., camel) is recognised and conceptualised, the phonological stage is thought to involve retrieval of phonological representations, often referred to as “word form retrieval”, and phonological encoding (e.g., /ˈkæməl/ with phonological segmentation and syllabification) before speech occurs. Breakdowns in naming can occur at any stage for various reasons and there has been reasonable assumption that in lvPPA naming deficits reflect a general phonological problem.^22,103^ Results from our previous study showed that lvPPA patients have impaired semantic performance and, in turn, we found that the semantic deficit positively correlated with naming scores.^47^ Of note, the nature of the semantic deficits in lvPPA is different from that in svPPA/SD, where naming impairments reflect degrading semantic representations due to bilateral atrophy centred on the anterior temporal lobes.^3,104,105^ Instead, there are commonalities in naming as well as other features of semantic control, between patients with lvPPA and those with post-stroke semantic aphasia and Wernicke’s aphasia.^48–53^ Despite the different aetiologies, the locus of damage is largely shared between semantic aphasia, Wernicke’s aphasia, and lvPPA, centred on the left posterior lateral temporal and temporoparietal junction.^9,106–108^ The posterior lateral temporal cortex and the temporoparietal junction cover areas that are implicated in semantic control and information buffering,^99,109,110^ and thus may explain why these deficits co-occur in lvPPA, post-stroke semantic aphasia and Wernicke’s aphasic patients.

### Is repetition and short-term memory performance related to the left hemisphere regions of interest?

Prior research strongly suggests that non-word repetition involves the pSTG, IFG, and premotor regions that are important in converting auditory speech sounds into articulatory-based representations.^59,111,112^ As expected in the current study, grey matter volumes were substantially reduced in the left pSTG, IFG, and ATL in the patients compared to controls. For the left dPM region, there was little or no evidence for differences between patients and controls, but this is an expected finding as the dPM/precentral gyrus is not a region typically atrophied in lvPPA and tAD patients. Likewise, we did not find any group differences in the levels of resting state functional connectivity between the four ROIs. In other words, even though patient groups showed significantly reduced grey matter volumes in three target brain regions, there were no changes in their functional connectivity. Our volumetric findings are consistent with previous studies reporting significant associations between phonological short-term memory tasks and atrophy in the left pSTG and IFG.^113–116^ In contrast to our connectivity findings, other studies have found either decreases or increases in functional connectivity in PPA, as well as other subtypes of FTD and AD.^116–121^ One possible explanation is that the patients in our sample were further on in their disease course compared to the presymptomatic or prodromal cases in the literature who can exhibit hyperconnectivity.^122^

When examining the associations between non-word repetition performance and grey matter volumes and functional connectivity between all ROIs, we found either no evidence or evidence for no relationship in the whole group. When excluding controls, the only anecdotal evidence that was corroborated by frequentist multiple regression was for an association between non-word repetition and pSTG-dPM functional connectivity after controlling for the grey matter volumes in the pSTG and dPM. The dorsal premotor cortex has been previously reported to be linked to phonological output with numerous studies finding its associations with non-word reading and repetition across patients with lvPPA, stroke aphasia, and healthy controls.^18,38,62,123^ In contrast, we found clear associations in the whole group between ACE-R multisyllabic word/phrase repetition and grey matter volumes in the pSTG, ATL and IFG. These findings replicate previous findings reporting significant positive correlations between atrophy in the pSTG and sentence repetition scores.^124,125^

### Limitations

There are limitations to our study. First, our sample size for the longitudinal patient data is relatively small. However, longitudinal investigations are rare in lvPPA samples, and we believe our data offer valuable insights regarding the trajectory of phonological and semantic decline in lvPPA. Second, the hearing loss status for each participant was based on the patient and family’s report. In the absence of standard audiometric testing or audiology assessment reports, we do not have further details regarding the degree of hearing loss for each participant. Given the small samples of hearing-impaired individuals and the lack of objective hearing measures, our study offers a tentative finding that requires further extension and explication. Finally, given the uncertainty regarding what the task-free fMRI precisely represents, our functional connectivity analyses were exploratory.

## Conclusion

We have shown that deficits in “pure” phonological tasks, namely single word and non-word repetition, are greater in patients with a self-reported hearing loss across all groups with lvPPA or tAD. Our detailed analyses of naming performance in lvPPA and how they relate to tasks of phonology, working memory, and semantics challenge the proposal that naming is solely due to impaired phonological processing in lvPPA. We propose that, instead of having a distinct core phonological impairment, lvPPA patients have a working memory/buffering-type impairment impacting their performance on length-dependent working memory tasks (e.g., sentence repetition).

## Data availability

The authors confirm that the data supporting the findings of this study are available within the article and its Supplementary material.

## Acknowledgements

We sincerely thank our patients and their families for supporting this work.

## Supplementary material

**Supplementary Table 1.**
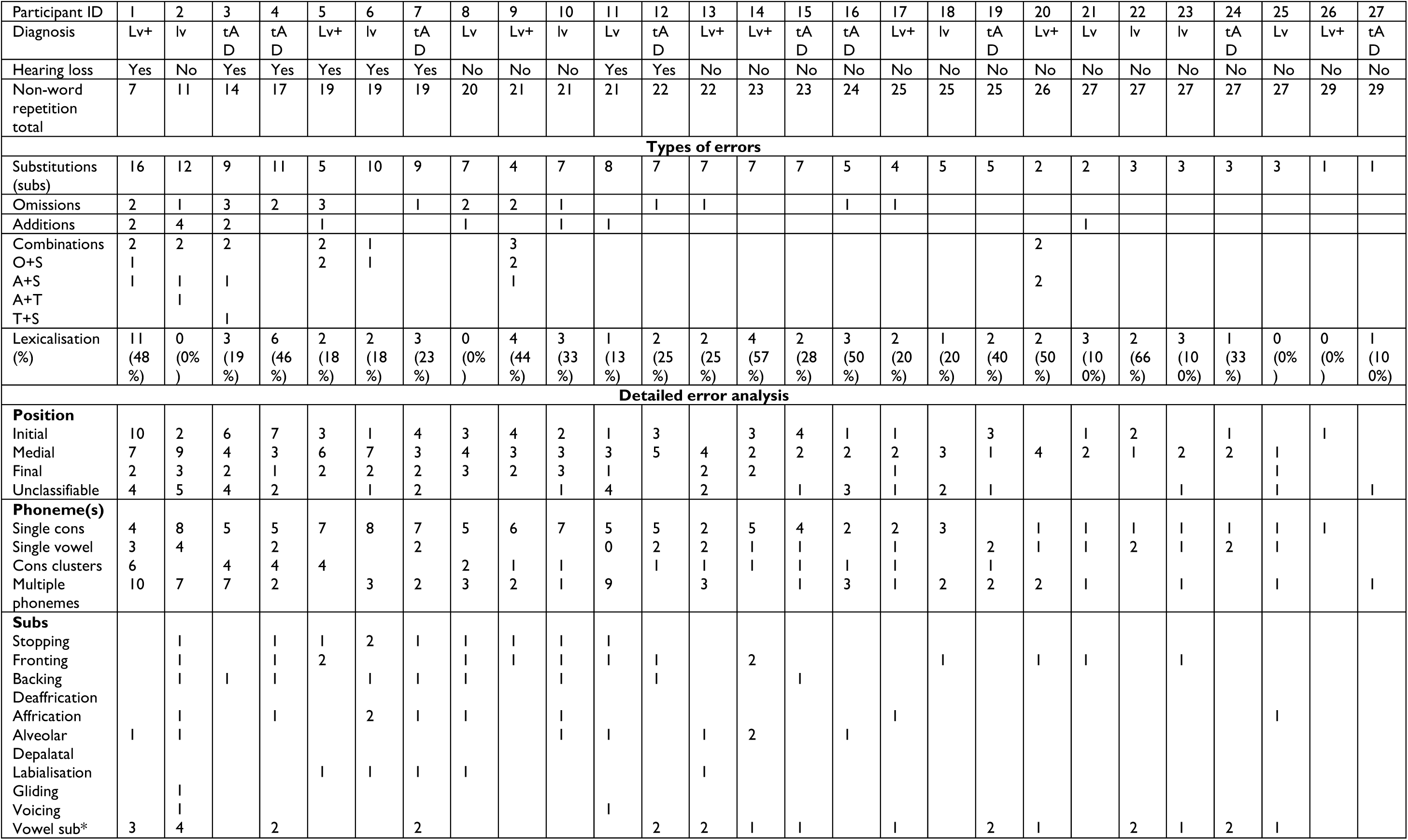
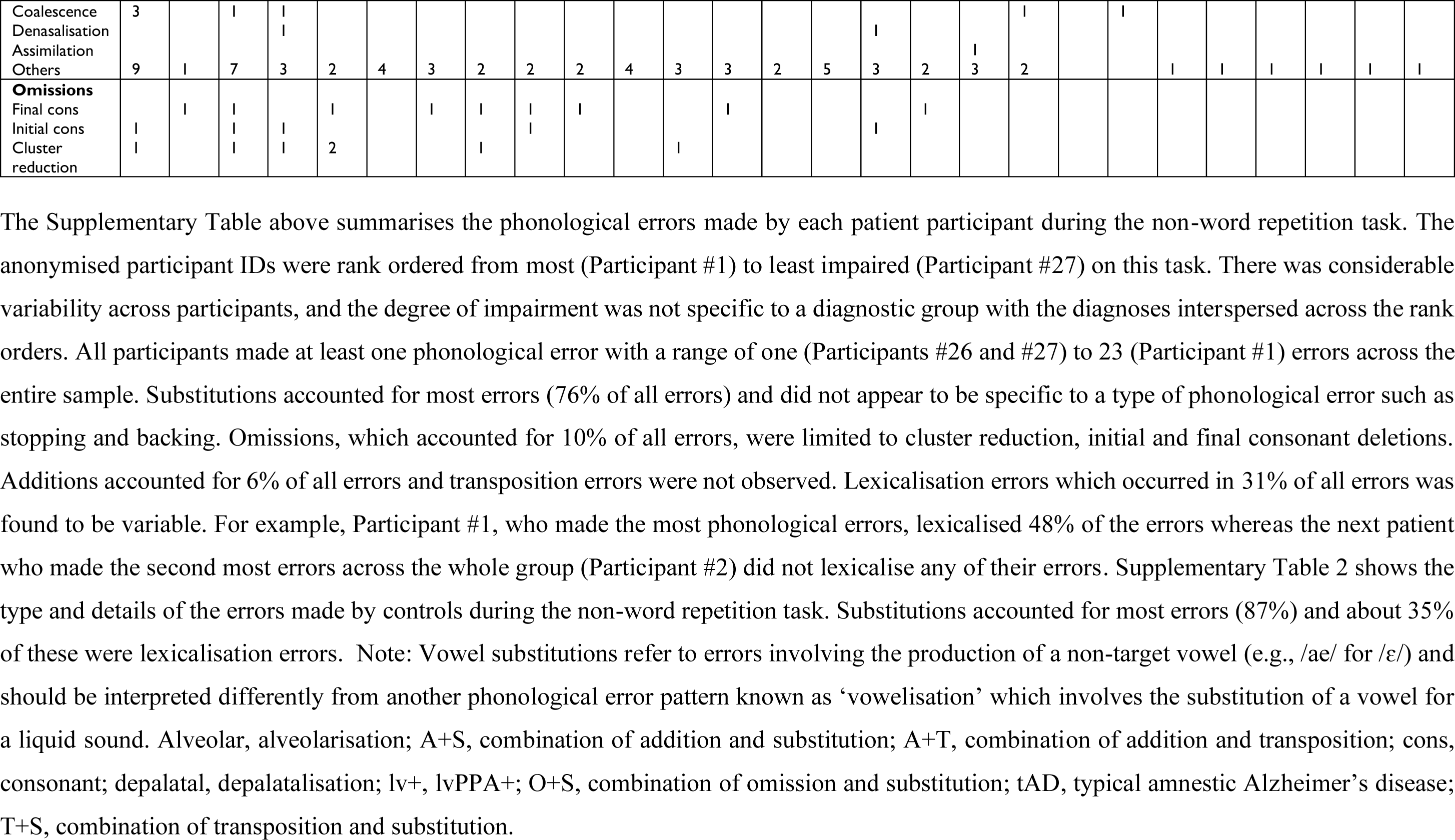
Detailed phonological error patterns in patients during PALPA non-word repetition.

**Supplementary Table 2.**
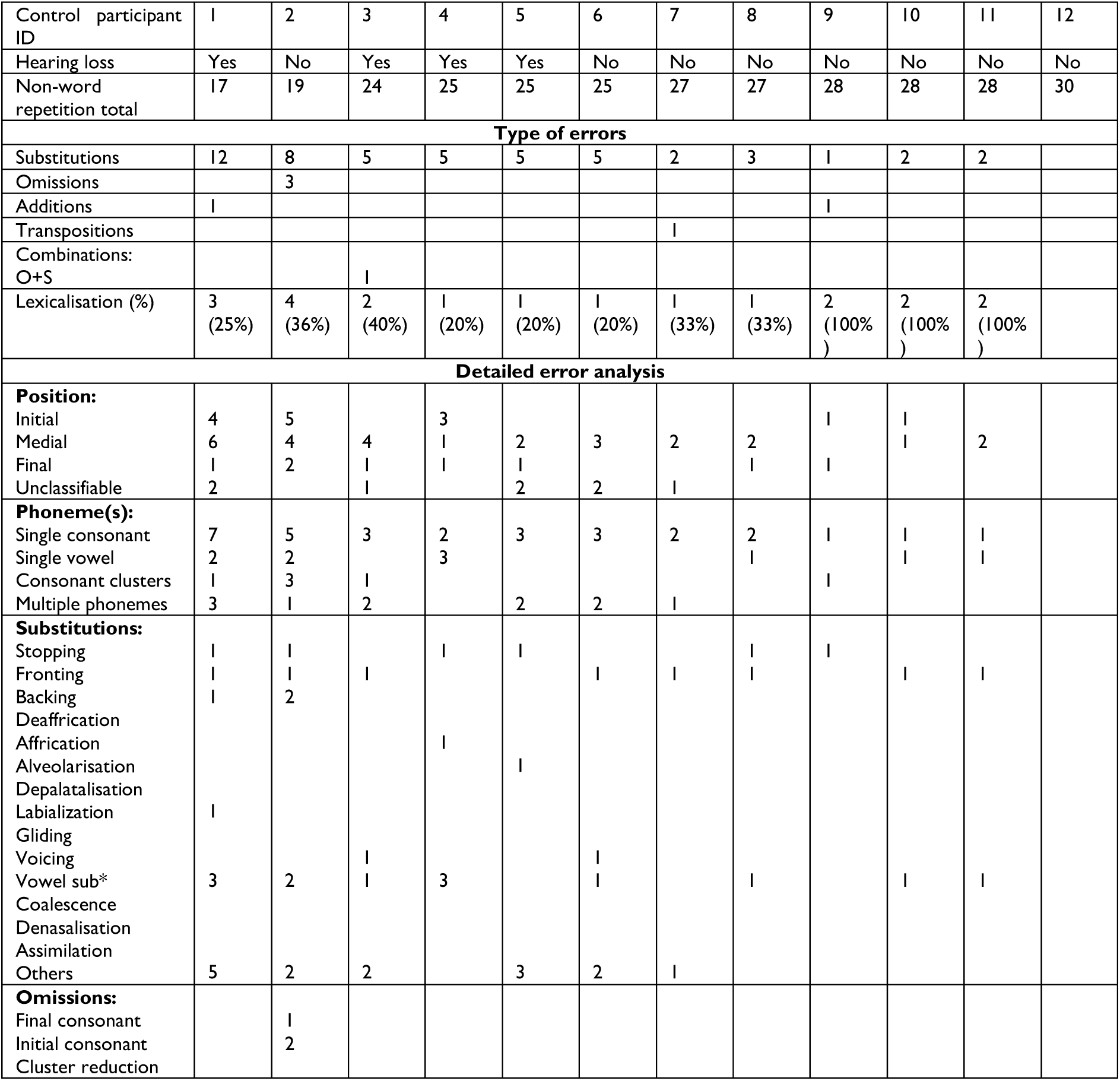
Detailed phonological error patterns in healthy controls during PALPA non-word repetition.

**Supplementary Table 3.** Bayesian correlations between non-word and multisyllabic word/phrase repetition and connected speech derived measures.

| Connected speech derived measure | All groups |  | tAD |  | lvPPA |  | lvPPA+ |  |
| --- | --- | --- | --- | --- | --- | --- | --- | --- |
|  | PALPA non-word repetition | ACE-R multisyllabic word/phrase repetition | PALPA non-word repetition | ACE-R multisyllabic word/phrase repetition | PALPA non-word repetition | ACE-R multisyllabic word/phrase repetition | PALPA non-word repetition | ACE-R multisyllabic word/phrase repetition |
| Phonological paraphasias per 100 words | $r = -0.24$, $P = 0.23$, BF = 20.48 | $r = -0.43$, $P = 0.03$, BF = 2.49 | $r = 0.21$, $P = 0.60$, BF = 0.46 | $r = 0.23$, $P = 0.55$, BF = 0.48 | $r = -0.25$, $P = 0.48$, BF = 0.48 | $r = -0.42$, $P = 0.23$, BF = 0.73 | **$r = -0.80$, $P = 0.02$, BF = 4.64** | $r = -0.24$, $P = 0.60$, BF = 0.52 |
| Fillers per 100 words | $r = 0.29$, $P = 0.14$, BF = 0.66 | $r = -0.27$, $P = 0.18$, BF = 0.58 | $r = 0.64$, $P = 0.07$, BF = 1.81 | $r = 0.47$, $P = 0.21$, BF = 0.82 | $r = 0.30$, $P = 0.39$, BF = 0.54 | $r = -0.41$, $P = 0.24$, BF = 0.72 | $r = 0.22$, $P = 0.61$, BF = 0.48 | **$r = 0.90$, $P = 0.006$, BF = 10.07** |
| Repaired sequences per 100 words | $r = -0.08$, $P = 0.71$, BF = 0.27 | $r = -0.45$, $P = 0.02$, BF = 2.97 | $r = 0.14$, $P = 0.72$, BF = 0.43 | $r = 0.06$, $P = 0.88$, BF = 0.41 | $r = 0.05$, $P = 0.90$, BF = 0.39 | $r = -0.44$, $P = 0.20$, BF = 0.80 | $r = -0.74$, $P = 0.04$, BF = 2.74 | $r = 0.53$, $P = 0.23$, BF = 0.87 |
| Pauses per 100 words | $r = -0.20$, $P = 0.33$, BF = 0.38 | **$r = -0.46$, $P = 0.02$, BF = 3.28** | $r = -0.47$, $P = 0.20$, BF = 0.84 | $r = 0.09$, $P = 0.81$, BF = 0.42 | $r = -0.16$, $P = 0.65$, BF = 0.42 | $r = -0.53$, $P = 0.12$, BF = 1.17 | $r = -0.36$, $P = 0.39$, BF = 0.60 | $r = 0.42$, $P = 0.35$, BF = 0.67 |
| Speech to pause ratio | $r = 0.35$, $P = 0.07$, BF = 1.09 | $r = 0.09$, $P = 0.65$, BF = 0.27 | $r = 0.46$, $P = 0.21$, BF = 0.81 | $r = -0.04$, $P = 0.92$, BF = 0.41 | $r = 0.34$, $P = 0.33$, BF = 0.59 | $r = 0.21$, $P = 0.57$, BF = 0.45 | $r = 0.37$, $P = 0.36$, BF = 0.62 | $r = -0.34$, $P = 0.46$, BF = 0.58 |
| Words to non-word utterances ratio | $r = 0.004$, $P = 0.99$, BF = 0.24 | $r = 0.29$, $P = 0.15$, BF = 0.64 | $r = -0.70$, $P = 0.04$, BF = 2.59 | $r = -0.30$, $P = 0.43$, BF = 0.53 | $r = 0.02$, $P = 0.97$, BF = 0.39 | **$r = 0.73$, $P = 0.02$, BF = 4.66** | $r = 0.60$, $P = 0.12$, BF = 1.24 | $r = -0.57$, $P = 0.19$, BF = 0.97 |
| Words per minute | $r = 0.09$, $P = 0.65$, BF = 0.26 | $r = 0.31$, $P = 0.12$, BF = 0.75 | $r = 0.38$, $P = 0.31$, BF = 0.64 | $r = 0.34$, $P = 0.37$, BF = 0.58 | $r = -0.29$, $P = 0.42$, BF = 0.52 | $r = 0.17$, $P = 0.63$, BF = 0.43 | $r = -0.38$, $P = 0.36$, BF = 0.62 | $r = -0.49$, $P = 0.27$, BF = 0.78 |
| Total utterances per minute | $r = -0.01$, $P = 0.97$, BF = 0.24 | $r = -0.06$, $P = 0.77$, BF = 0.25 | **$r = 0.73$, $P = 0.03$, BF = 3.43** | $r = 0.23$, $P = 0.56$, BF = 0.47 | $r = -0.37$, $P = 0.30$, BF = 0.63 | $r = -0.25$, $P = 0.49$, BF = 0.48 | $r = -0.07$, $P = 0.87$, BF = 0.43 | $r = -0.32$, $P = 0.49$, BF = 0.56 |
| Fillers per minute | $r = 0.37$, $P = 0.06$, BF = 1.26 | $r = -0.03$, $P = 0.90$, BF = 0.25 | $r = 0.59$, $P = 0.10$, BF = 1.35 | $r = 0.47$, $P = 0.21$, BF = 0.83 | $r = 0.28$, $P = 0.43$, BF = 0.51 | $r = -0.28$, $P = 0.43$, BF = 0.51 | $r = 0.37$, $P = 0.37$, BF = 0.61 | $r = 0.68$, $P = 0.09$, BF = 1.54 |
| Pauses per minute | **$r = -0.44$, $P = 0.02$, BF = 3.10** | $r = -0.29$, $P = 0.15$, BF = 0.66 | $r = -0.31$, $P = 0.42$, BF = 0.54 | $r = 0.42$, $P = 0.26$, BF = 0.71 | $r = -0.66$, $P = 0.04$, BF = 2.54 | $r = -0.47$, $P = 0.17$, BF = 0.90 | $r = -0.44$, $P = 0.28$, BF = 0.72 | $r = 0.25$, $P = 0.59$, BF = 0.52 |
| Long (>2 seconds) pauses per minute | $r = -0.216$, $P = 0.20$, BF = 0.53 | $r = -0.18$, $P = 0.38$, BF = 0.35 | $r = -0.49$, $P = 0.18$, BF = 0.88 | $r = -0.02$, $P = 0.95$, BF = 0.41 | $r = -0.39$, $P = 0.27$, BF = 0.66 | $r = -0.08$, $P = 0.83$, BF = 0.39 | $r = -0.03$, $P = 0.95$, BF = 0.43 | $r = 0.05$, $P = 0.92$, BF = 0.46 |
| Short pauses (<2 seconds) per minute | $r = -0.34$, $P = 0.08$, BF = 1.03 | $r = -0.22$, $P = 0.27$, BF = 0.43 | $r = 0.07$, $P = 0.85$, BF = 0.41 | $r = 0.63$, $P = 0.07$, BF = 1.76 | $r = -0.42$, $P = 0.22$, BF = 0.75 | $r = -0.46$, $P = 0.18$, BF = 0.87 | $r = -0.65$, $P = 0.08$, BF = 1.63 | $r = 0.42$, $P = 0.36$, BF = 0.67 |
| Repaired sequences per minute | $r = -0.13$, $P = 0.51$, BF = 0.29 | $r = -0.35$, $P = 0.08$, BF = 1.02 | $r = 0.24$, $P = 0.53$, BF = 0.48 | $r = 0.15$, $P = 0.69$, BF = 0.44 | $r = 0.04$, $P = 0.91$, BF = 0.39 | $r = -0.38$, $P = 0.28$, BF = 0.65 | $r = -0.64$, $P = 0.09$, BF = 1.55 | $r = 0.24$, $P = 0.60$, BF = 0.52 |

Note: Bayes factor greater than 3 and their associated Pearson's correlation values are bolded. ACE-R, Addenbrooke's Cognitive Examination – Revised; lvPPA, logopenic variant of primary progressive aphasia; PALPA, the Psycholinguistic Assessment of Language Processing in Aphasia; tAD, typical Alzheimer's disease.

**Supplementary Table 4.**
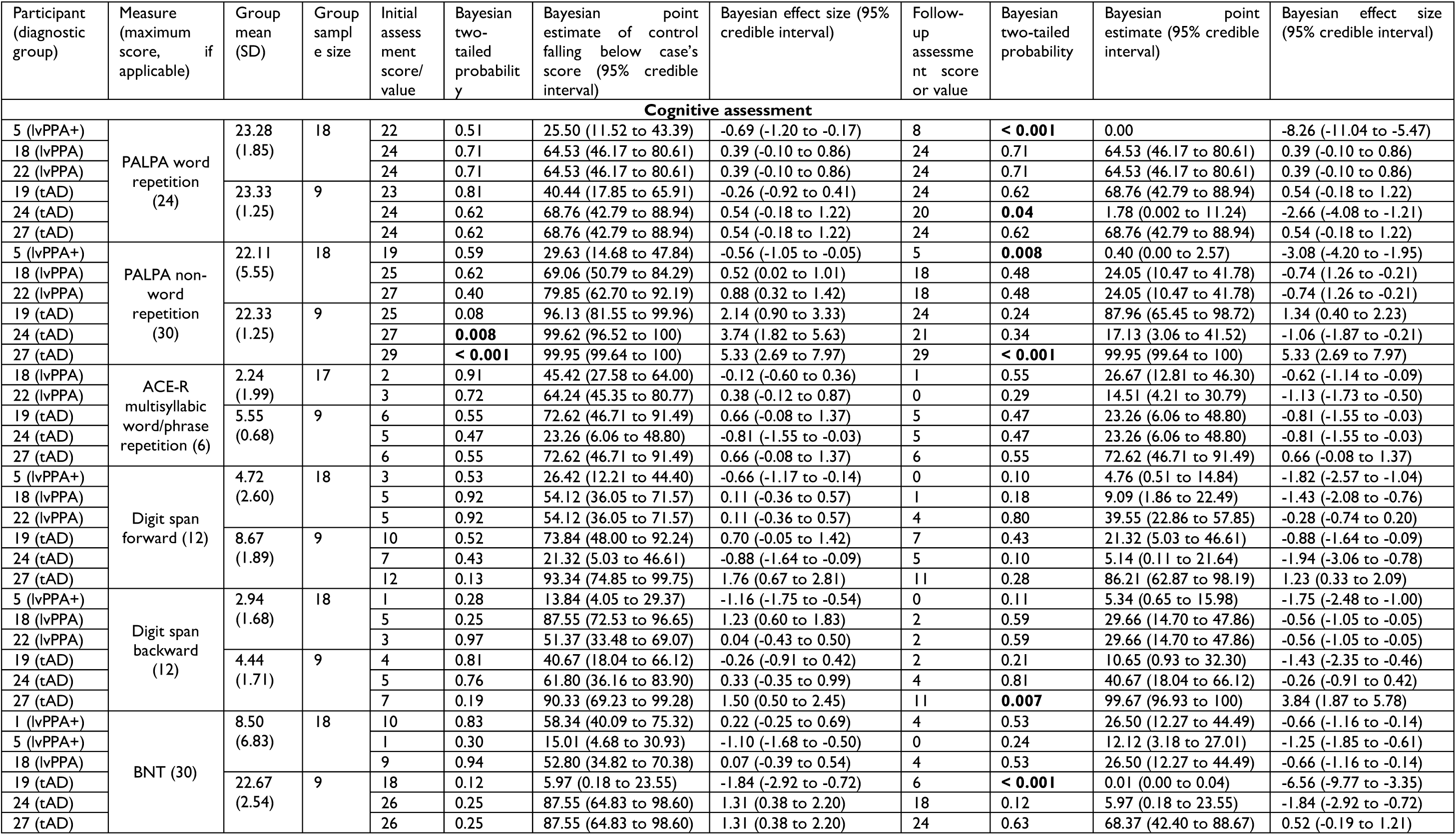
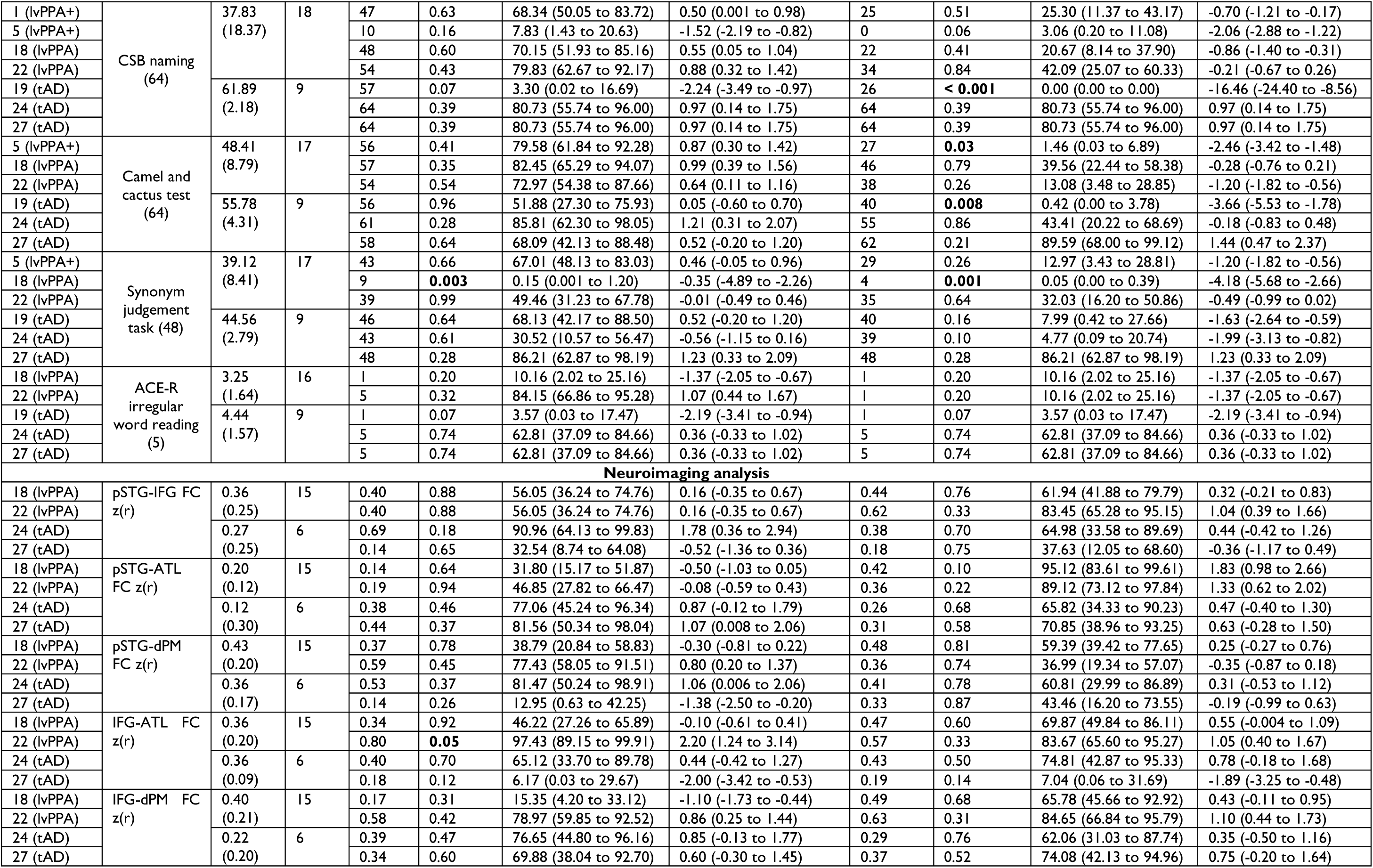
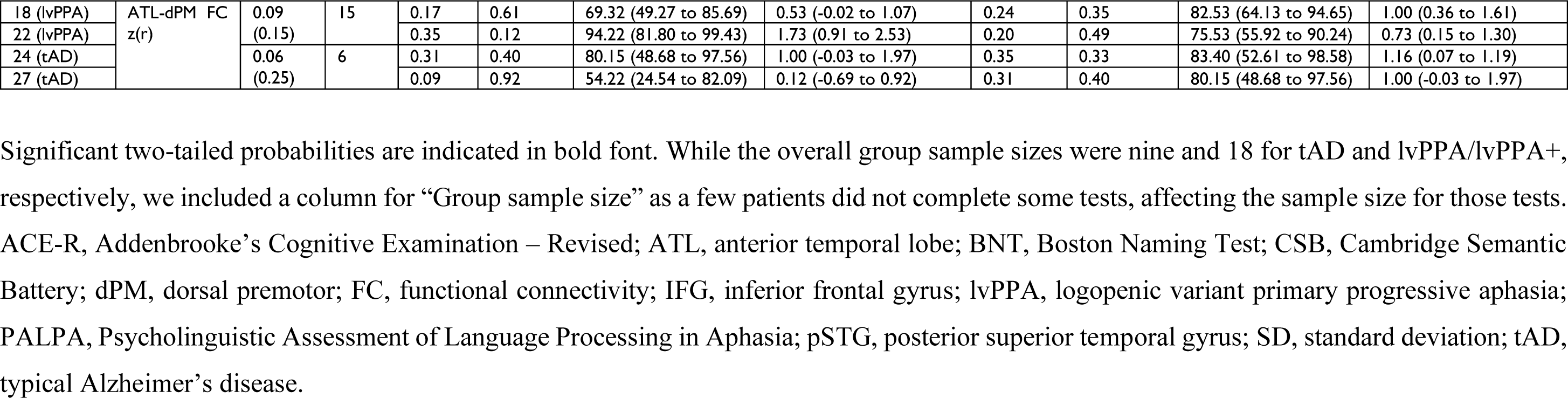
Bayesian point and interval estimates of effect sizes for each longitudinal case relative to their respective diagnostic group sample.

**Supplementary Figure 1.**
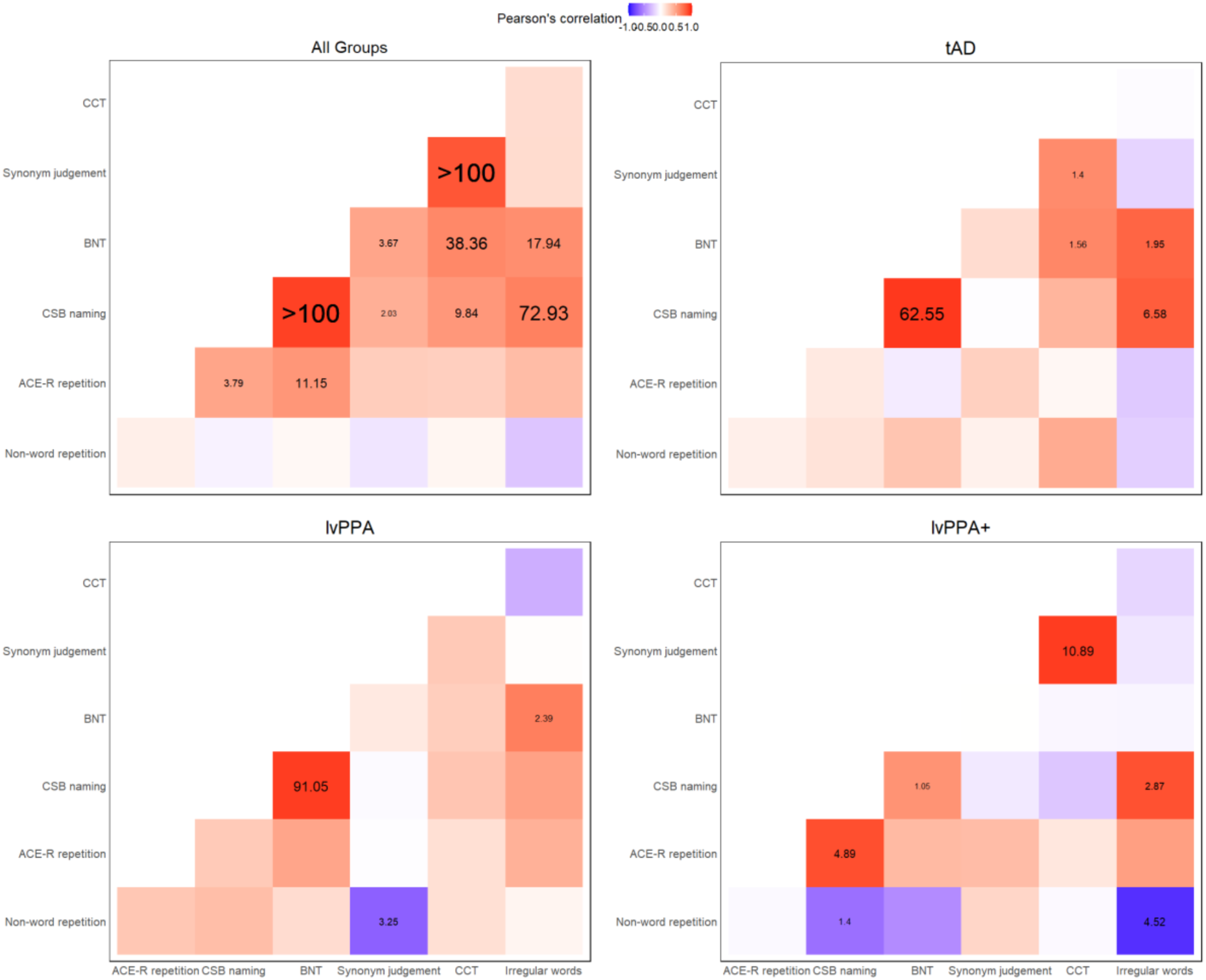
Associations between PALPA non-word and ACE-R multisyllabic word/phrase repetition and semantic assessments. Pearson’s correlation values are represented in the colour legend, where red and purple colours represent positive and negative correlations, respectively. The associated Bayes factors (BFs) are shown in black text, where the larger versus smaller fonts denote the degree of evidence (e.g., extreme for BF >100 versus anecdotal for 1 < BF < 3). In the individual patient groups, the only other moderate evidence was found between non-word repetition and synonym judgement task in the lvPPA group (*r* = -0.69, *P* = 0.03, BF = 3.25), as well as between non-word repetition and ACE-R irregular word reading (*r* = -0.89, *P* = 0.02, BF = 4.52) and between ACE-R multisyllabic word/phrase repetition and CBS naming (*r* = 0.84, *P* = 0.04, BF = 4.89) in the lvPPA+ group.

**Supplementary Figure 2.**
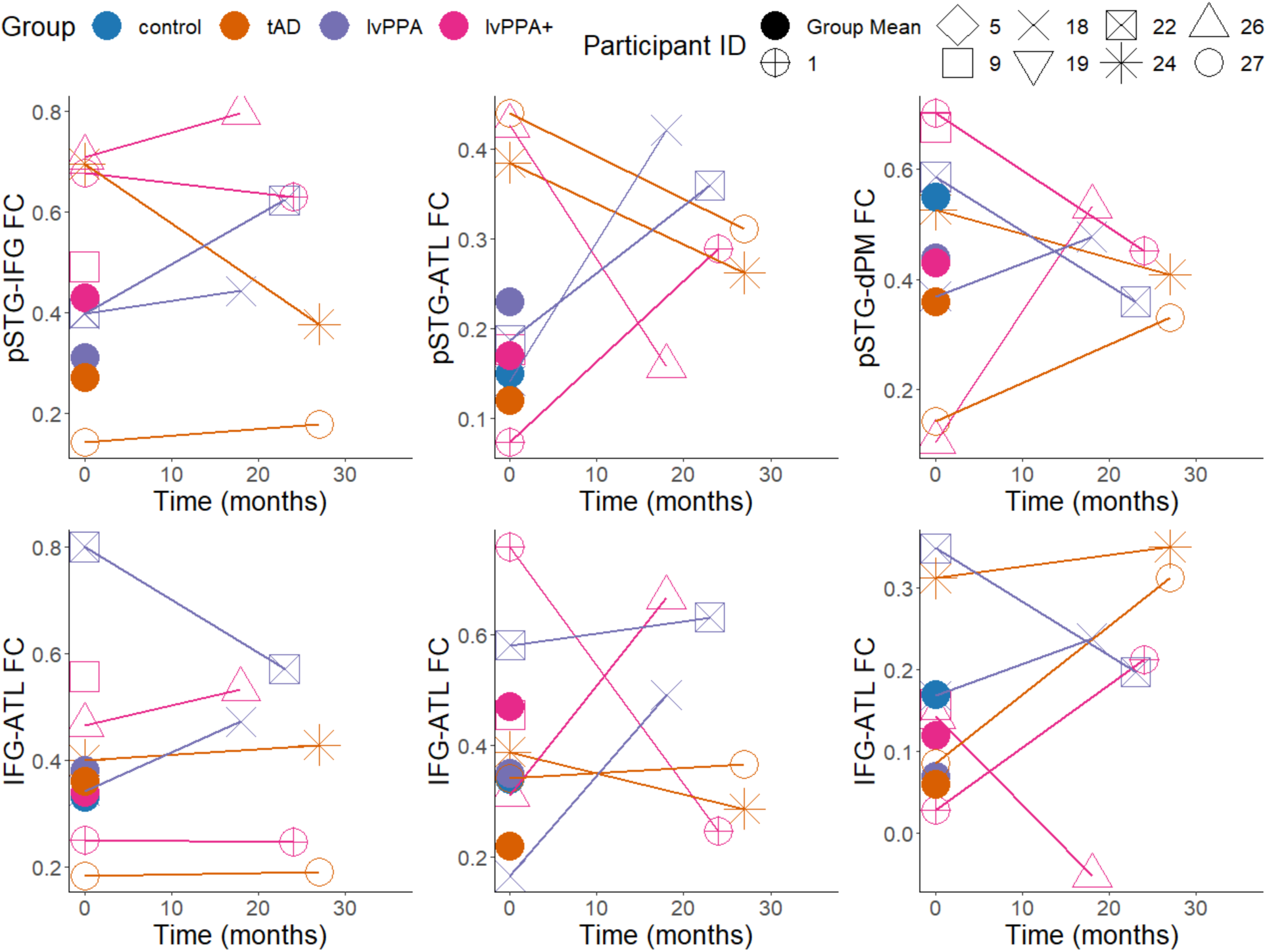
**Functional connectivity z-values between all region-of-interests (ROIs) over time in six patient participants**. The x-axis represents the time from to initial to follow-up assessment in months. The filled colour dots represent the group average functional connectivity z-values for each ROI pair. The connected lines indicate the scores of each participant who completed the same test at two time-points. Note: The lines merely connect the same participants across two time-points and do not suggest a linear decline over time. ATL, anterior temporal lobe; dPM, dorsal premotor region; FC, functional connectivity; IFG, inferior frontal gyrus, lvPPA, logopenic variant of primary progressive aphasia; pSTG, posterior superior temporal gyrus; tAD, typical Alzheimer’s disease.

